# Biallelic null variants in *PNPLA8* cause microcephaly through the reduced abundance of basal radial glia

**DOI:** 10.1101/2023.04.26.23288947

**Authors:** Yuji Nakamura, Issei S. Shimada, Reza Maroofian, Henry Houlden, Micol Falabella, Masanori Fujimoto, Emi Sato, Hiroshi Takase, Shiho Aoki, Akihiko Miyauchi, Eriko Koshimizu, Satoko Miyatake, Yuko Arioka, Mizuki Honda, Takayoshi Higashi, Fuyuki Miya, Yukimune Okubo, Isamu Ogawa, Annarita Scardamaglia, Mohammad Miryounesi, Sahar Alijanpour, Farzad Ahmadabadi, Peter Herkenrath, Hormos Salimi Dafsari, Clara Velmans, Mohammed Balwi, Antonio Vitobello, Anne-Sophie Denommé-Pichon, Médéric Jeanne, Antoine Civit, Maha S. Zaki, Hossein Darvish, Somayeh Bakhtiari, Michael Kruer, Christopher J Carroll, Ehsan Ghayoor Karimiani, Rozhgar A Khailany, Talib Adil Abdulqadir, Mehmet Ozaslan, Peter Bauer, Giovanni Zifarelli, Tahere Seifi, Mina Zamani, Chadi Al Alam, Robert D S Pitceathly, Kazuhiro Haginoya, Tamihide Matsunaga, Hitoshi Osaka, Naomichi Matsumoto, Norio Ozaki, Yasuyuki Ohkawa, Shinya Oki, Tatsuhiko Tsunoda, Yoshitaka Taketomi, Makoto Murakami, Yoichi Kato, Shinji Saitoh

**Author notes:** Correspondence to: Issei S Shimada, Yoichi Kato, and Shinji Saitoh Issei S. Shimada: Department of Cell Biology, Nagoya City University Graduate School of Medical Sciences, Kawasumi 1, Mizuho-cho, Mizuho-ku, Nagoya, Aichi 467-8601, Japan; E- mail; Yoichi Kato: Department of Cell Biology, Nagoya City University Graduate School of Medical Sciences, Kawasumi 1, Mizuho-cho, Mizuho-ku, Nagoya, Aichi 467-8601, Japan; Shinji Saitoh: Department of Pediatrics and Neonatology, Nagoya City University Graduate School of Medical Sciences, Kawasumi 1, Mizuho-cho, Mizuho-ku, Nagoya 467-8601, Japan. **Authorship note:** ISS, YK, and SS are co-senior authors. **Conflict of interest:** The authors have declared that no conflict of interest exists.

## Abstract

PNPLA8, one of the calcium-independent phospholipase A2 enzymes, is involved in various physiological processes through the maintenance of membrane phospholipids. However, little is known about its role in brain development. Here, we report 12 individuals from 10 unrelated families with biallelic ultra-rare variants in *PNPLA8* presenting with a wide spectrum of clinical features ranging from developmental and epileptic-dyskinetic encephalopathy (DEDE) to progressive movement disorders. Complete loss of PNPLA8 was associated with the severe end of the spectrum, showing DEDE manifestations and congenital or progressive microcephaly. Using cerebral organoids generated from human induced pluripotent stem cells, we found that loss of PNPLA8 reduced the number of basal radial glial cells (bRGCs) and upper-layer neurons. By spatial transcriptomic analysis targeting apical radial glial cells (aRGCs), we found the downregulation of bRGC-related gene sets in patient-derived cerebral organoids. Lipidomic analysis revealed a decrease in the amount of lysophosphatidic acid, lysophosphatidylethanolamine, and phosphatidic acid, indicative of the disturbed phospholipid metabolism in *PNPLA8* knockout neural progenitor cells. Our data suggest that PNPLA8 has a critical role in the bRGC-mediated expansion of the developing human cortex by regulating the fate commitment of aRGCs.

## Introduction

In the evolutionary expanded cerebral cortex, the increased proliferation of neural stem and progenitor cells (NPCs) underlies the increased number of neurons (1). Basal radial glial cells (bRGCs) are a distinct type of NPC and are characterized by a significant proliferative potential compared with the other types of NPCs (2–4). bRGCs originate from apical radial glial cells (aRGCs). When aRGCs divide, they produce diverse cell types other than bRGCs, such as aRGCs, neurons, and basal intermediate progenitor cells (bIPs) (5). In contrast, most bRGCs undergo self-renewal divisions, hence leading to exponential amplification (2) and the abundance of bRGCs has been linked to the highly expanded and folded cerebral cortex (3).

Patatin-like phospholipase domain-containing lipase 8 (PNPLA8), also called calcium-independent phospholipase A2γ, is a phospholipase A2 (PLA2) enzyme conserved across species (6–8). *PNPLA8* transcripts have multiple translation initiation sites, thereby expressing multiple protein sizes (MWs of 88, 77, 74, and 63 kDa) (9, 10). PNPLA8 was shown to be expressed in distinct tissues, including the brain (11). The subcellular localization of PNPLA8 is unique in that it has a mitochondrial localization sequence unlike other PNPLA family members sharing the common lipase consensus domain (10). PNPLA8 hydrolyzes phospholipids to generate lysophospholipids and free fatty acids (FFAs), thereby regulating multiple cellular processes, namely membrane remodeling (12–14), maintenance of mitochondrial function (12, 13, 15), protection against oxidative stress (16, 17), and lipid mediator biosynthesis (18, 19).

The loss of PNPLA8 function affects the central nervous system in mice and humans (12, 13, 20–23). Neurodegenerative phenotypes were documented in *Pnpla8* KO mice as they exhibited spatial learning and memory deficits with morphologic alterations of mitochondria in the hippocampus despite the gross anatomy of their brains appearing normal (12). Biallelic variants in *PNPLA8* in humans were initially reported to cause neurodegeneration and/or myopathy as observed in *Pnpla8* KO mouse models (22). However, additional reports have raised the possibility of wider phenotypic variability since some patients showed early-onset neurological manifestations accompanied by brain malformations (20–23). However, due to the small number of affected individuals, the clinical features have not been fully detailed, and the possible occurrence of brain malformations underscores the importance of elucidating unknown roles for PNPLA8 in human brain development.

Here, we report 14 individuals from 12 unrelated families with biallelic variants in *PNPLA8*. We describe a human cerebral organoid model lacking *PNPLA8* to investigate the loss-of-function (LoF) nature of *PNPLA8* variants specifically in cortical development. We further explore the disease mechanism with multi-omics approaches. Our data detail the clinical features associated with the type of *PNPLA8* variants and indicate that a complete loss of PNPLA8 impairs bRGC-mediated cortical expansion in humans.

## Results

### General characterization of biallelic variants in PNPLA8

We identified 12 individuals showing neurological manifestations from 10 unrelated families with biallelic variants in *PNPLA8* (Figure 1A). The age of the patients at the last assessment ranged (removed due to medRxiv policy) of age (Table 1). We identified 12 variants, of which 10 were novel. Of the 12 variants, four were nonsense, four were frameshift, three were intronic variants near an exon-intron boundary, and one was a missense variant. According to the population-based variant frequency data, such as Single Nucleotide Polymorphism database (24), 1000Genomes (25), jMorp (26), and Genome Aggregation Database (27), the frequency of all of the variants was less than 0.001%, suggesting a strong purification effect due to disease pathogenicity.

**Figure 1.**
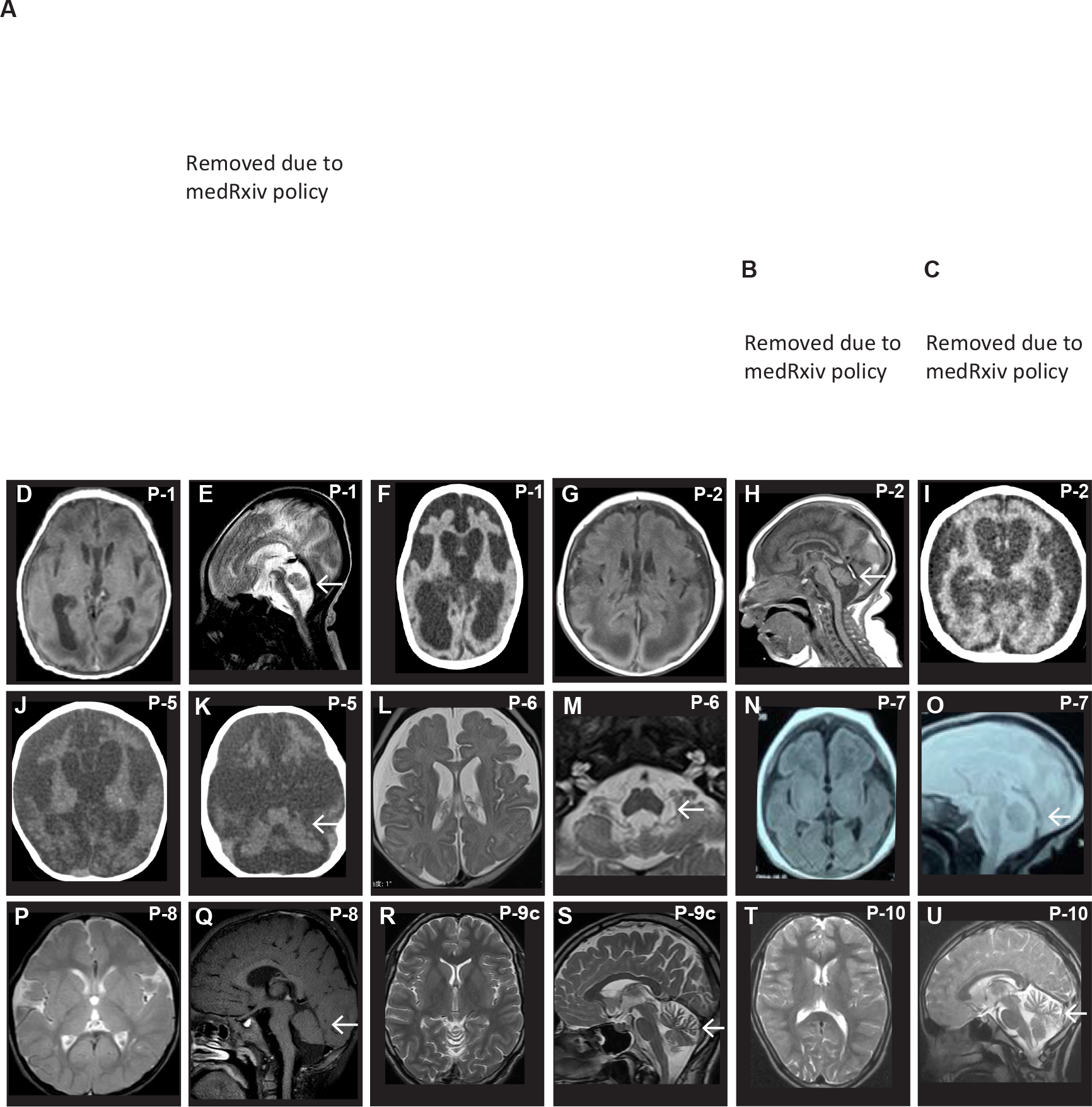
Identification of biallelic loss-of-function variants in *PNPLA8* in patients with diverse neurological phenotypes. (**A**) Pedigrees showing autosomal recessive inheritance with biallelic *PNPLA* variants in 10 unrelated families. The probands are indicated by arrows. Black fills denote affected individuals, squares represent males, and circles represent females. Double lines indicate first cousin status. Pathogenic or likely pathogenic variants in *PNPLA8* are denoted as ‘mut’, and WT sequences in *PNPLA8* are represented as ‘wt’. (**B** and **C**) Clinical photographs of Patient 1 and 7. (**D**-**F**) Axial T1-weighted (**D**) and sagittal T2-weighted (**E**) MRI images and a brain CT image (**F**) of Patient 1. The arrow indicates pontocerebellar hypoplasia. (**G**-**I**) T1-weighted images of MRI (**G** and **H**) and a brain CT image (**I**) of Patient 2. The arrow indicates pontocerebellar hypoplasia. (**J** and **K**) Brain CT images of Patient 5. The arrow indicates cerebellar atrophy. (**L** and **M**) T2-weighted MRI images of Patient 6. The arrow indicates pontocerebellar hypoplasia. (**N** and **O**) Axial T1-weighted (**N**) and sagittal T2- weighted (**O**) MRI images of Patient 7. The arrow indicates pontocerebellar hypoplasia. (**P** and **Q**) Axial T2-weighted (**P**) and sagittal T1-weighted (**Q**) MRI images of Patient 8. The arrow indicates pontocerebellar hypoplasia. (**R** and **S**) T2-weighted MRI images of Patient 9c, exhibiting cerebellar atrophy (arrow). (**T** and **U**) T2-weighted MRI images of Patient 10, exhibiting cerebellar atrophy (arrow).

**Table 1.**
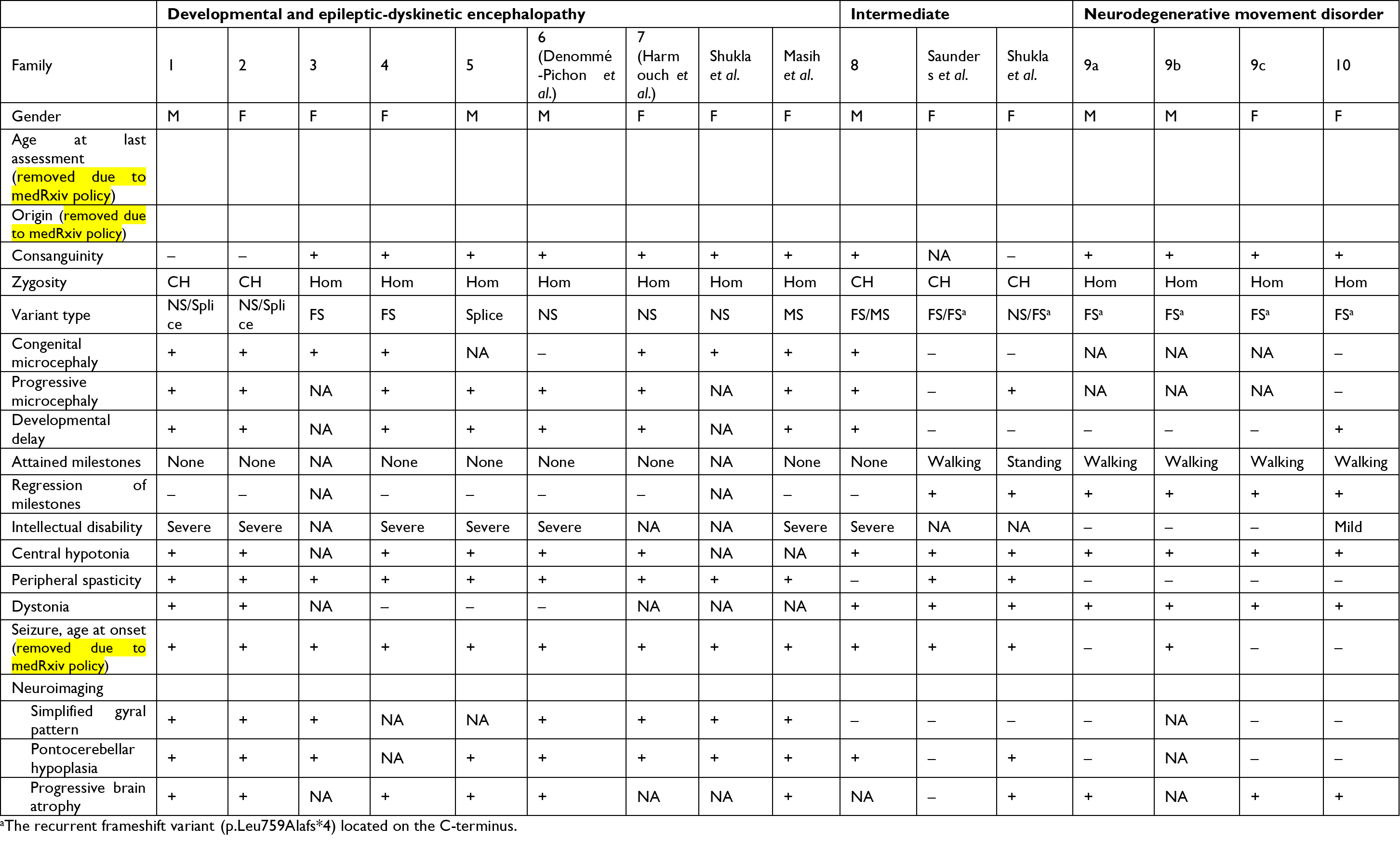
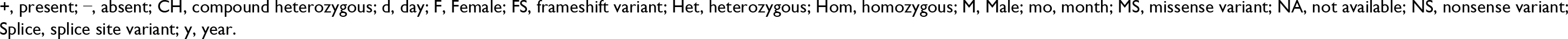
Summary of clinical features for affected individuals with *PNPLA8* variants

|  | Developmental and epileptic-dyskinetic encephalopathy |  |  |  |  |  |  |  |  | Intermediate |  |  | Neurodegenerative movement disorder |  |  |  |
| --- | --- | --- | --- | --- | --- | --- | --- | --- | --- | --- | --- | --- | --- | --- | --- | --- |
| Family | 1 | 2 | 3 | 4 | 5 | 6<br>(Denommé<br>-Pichon et<br>al.) | 7<br>(Harmouch<br>et al.) | Shukla<br>et al. | Masih<br>et al. | 8 | Saunders<br>et al. | Shukla<br>et al. | 9a | 9b | 9c | 10 |
| Gender | M | F | F | F | M | M | F | F | F | M | F | F | M | M | F | F |
| Age at last<br>assessment<br>(removed due to<br>medRxiv policy) |  |  |  |  |  |  |  |  |  |  |  |  |  |  |  |  |
| Origin (removed due<br>to medRxiv policy) |  |  |  |  |  |  |  |  |  |  |  |  |  |  |  |  |
| Consanguinity | – | – | + | + | + | + | + | + | + | + | NA | – | + | + | + | + |
| Zygosity | CH | CH | Hom | Hom | Hom | Hom | Hom | Hom | Hom | CH | CH | CH | Hom | Hom | Hom | Hom |
| Variant type | NS/Splice | NS/Splice | FS | FS | Splice | NS | NS | NS | MS | FS/MS | FS/FS<sup>a</sup> | NS/FS<sup>a</sup> | FS<sup>a</sup> | FS<sup>a</sup> | FS<sup>a</sup> | FS<sup>a</sup> |
| Congenital<br>microcephaly | + | + | + | + | NA | – | + | + | + | + | – | – | NA | NA | NA | – |
| Progressive<br>microcephaly | + | + | NA | + | + | + | + | NA | + | + | – | + | NA | NA | NA | – |
| Developmental<br>delay | + | + | NA | + | + | + | + | NA | + | + | – | – | – | – | – | + |
| Attained milestones | None | None | NA | None | None | None | None | NA | None | None | Walking | Standing | Walking | Walking | Walking | Walking |
| Regression of<br>milestones | – | – | NA | – | – | – | – | NA | – | – | + | + | + | + | + | + |
| Intellectual disability | Severe | Severe | NA | Severe | Severe | Severe | NA | NA | Severe | Severe | NA | NA | – | – | – | Mild |
| Central hypotonia | + | + | NA | + | + | + | + | NA | NA | + | + | + | + | + | + | + |
| Peripheral spasticity | + | + | + | + | + | + | + | + | + | – | + | + | – | – | – | – |
| Dystonia | + | + | NA | – | – | – | NA | NA | NA | + | + | + | + | + | + | + |
| Seizure, age at onset<br>(removed due to<br>medRxiv policy) | + | + | + | + | + | + | + | + | + | + | + | + | – | + | – | – |
| Neuroimaging |  |  |  |  |  |  |  |  |  |  |  |  |  |  |  |  |
| Simplified gyral<br>pattern | + | + | + | NA | NA | + | + | + | + | – | – | – | – | NA | – | – |
| Pontocerebellar<br>hypoplasia | + | + | + | NA | + | + | + | + | + | + | – | + | – | NA | – | – |
| Progressive brain<br>atrophy | + | + | NA | + | + | + | NA | NA | + | NA | – | + | + | NA | + | + |

<sup>a</sup>The recurrent frameshift variant (p.Leu759Alafs\*4) located on the C-terminus.
+, present; −, absent; CH, compound heterozygous; d, day; F, Female; FS, frameshift variant; Het, heterozygous; Hom, homozygous; M, Male; mo, month; MS, missense variant; NA, not available; NS, nonsense variant; Splice, splice site variant; y, year.

Regarding the impact on splicing, c.1625+3_1625+6del identified in the patient from Family 1 (Patient 1) was assessed using patient blood-derived lymphoblastoid cell lines.

Sequencing of each separated transcript by TA cloning revealed that c.1625+3_1625+6del resulted in the skipping of exons 6-8 due to aberrant splicing (Supplemental Figure 1, A and B). Due to the lack of patient-derived cells, we could not experimentally assess the splicing impact of c.2075-2A>G in Patient 2 and c.1684-2A>G in Patient 5. However, *in silico* analysis with SpliceAI (28) predicted that c.2075-2A>G and c.1684-2A>G results in the loss of a splice acceptor site with a Δ score of 0.97 and 0.93, respectively (high specificity cutoff: 0.8). Collectively, 11 out of 12 individuals carried the combination of nonsense, frameshift, and splice-site variants, which suggests LoF in PNPLA8.

### Clinical spectrum of PNPLA8-related neurological disease

We collected clinical information from the newly identified patients (Table 1, additional details in Supplemental Table 1, case reports in the Supplemental Text). Highly frequent clinical features of our patients were central hypotonia (100%, 11/11), dystonia (70%, 7/10), seizure (75%, 9/12), and progressive brain atrophy identified by neuroimaging (100%, 8/8). Among the 12 patients, seven patients (Families 1-7) were further characterized by several common features, such as congenital microcephaly (83%, 5/6; Figure 1, B and C), global developmental delay attaining no milestones (100%, 6/6), peripheral spasticity (100%, 7/7), and early-onset seizure (removed due to medRxiv policy) (100%, 7/7). Neuroimaging revealed that these patients showed simplified gyral pattern (100%, 5/5), pontocerebellar hypoplasia (100%, 6/6), and severe progressive atrophy of the cerebral cortex (100%, 5/5; Figure 1, D-O). In contrast, four patients (Families 9-10) were characterized by the following common features; attaining the milestone of walking (100%, 4/4), ataxia (100%, 4/4), regression of milestones (100%, 4/4), mild or no intellectual disability (100%, 4/4), and unique or absent seizures (100%, 4/4). Brain MRI showed little or no cerebral cortical volume loss but progressive cerebellar atrophy, consistent with the pronounced movement disorder observed in patients (100%, 3/3; Figure 1, R-U, Supplemental Video 1 and 2). The remaining patient (Family 8) showed intermediate severity as he showed global developmental delay and congenital microcephaly but later onset seizures (removed due to medRxiv policy). Brain MRI also showed pontocerebellar hypoplasia (Figure 1, P and Q).

We next assessed the correlation between the phenotypic features and the corresponding *PNPLA8* variants. By combining previously reported cases, we performed intergroup comparisons (Table 1, Supplemental Table 1, Figure 2A). We found that the clinical features varied according to the variant types and were classified into three groups. Seven patients (Familes 1-7) and two previously reported cases were on the severe end of the spectrum, which we termed “developmental and epileptic-dyskinetic encephalopathy (DEDE)” (Table 1). All of these patients carried biallelic LoF variants affecting three or four PNPLA8 protein isoforms. (Figure 2A). One patient (Family 8) and two previously reported cases had intermediate severity (Table 1). These patients carried compound heterozygous variants; one was an LoF variant located across at least three protein isoforms, and the other was a missense variant or the recurrent frameshift variant (p.Leu759Alafs*4) located at the C-terminus (Figure 2A). Four patients (Families 9-10) were in the relatively mild group, which we termed “neurodegenerative movement disorder” (Table 1). These patients were homozygous for the recurrent C-terminal frameshift variant (Figure 2A).

**Figure 2.**
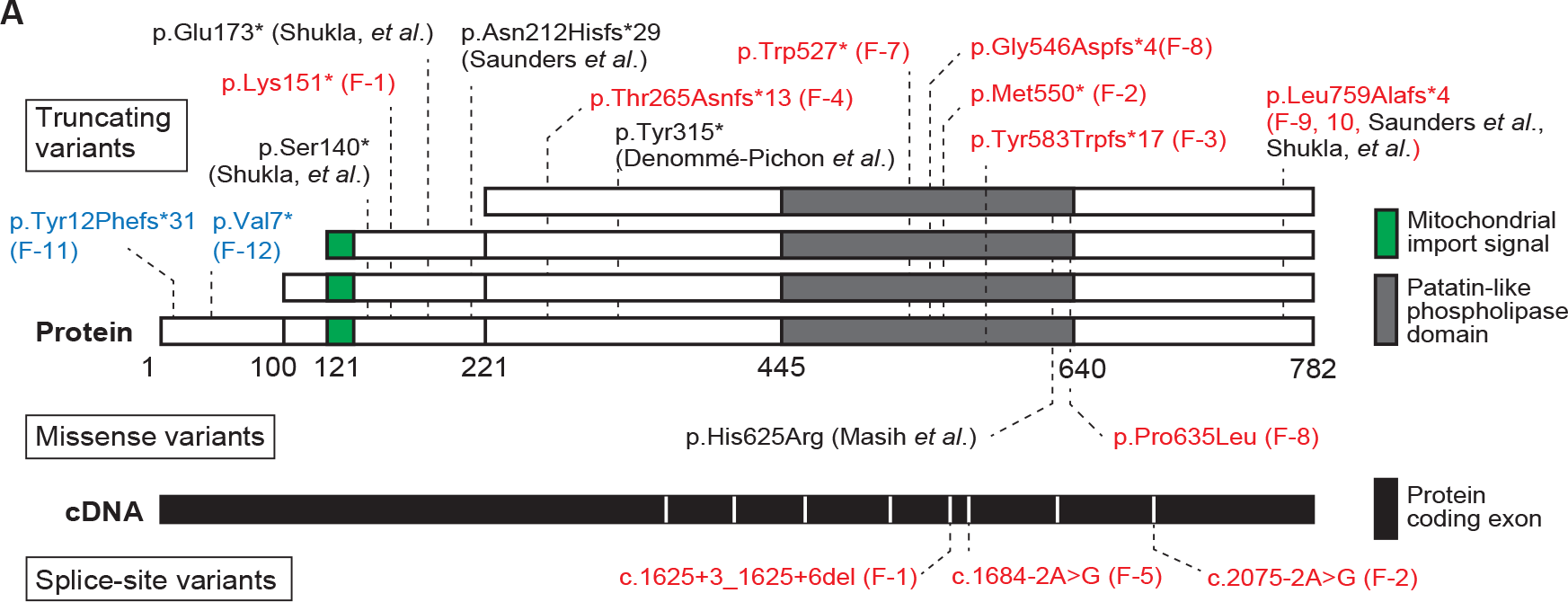
PNPLA8 protein isoforms with the position of the identified variants. (**A**) Schematic representation of PNPLA8 protein isoforms due to alternative translation initiation sites (*top*) and *PNPLA8* exon locations reflecting the canonical full-length transcript (NM_001256007.3, *bottom*). Disease-associated variants in this study (*red*) and previous studies (*black*) are noted. The identified variants from non-neuronal cohorts are noted in *blue*.

To evaluate the molecular effects of the identified variants, we obtained skin fibroblasts from Patient 1 (DEDE phenotype with biallelic LoF variants) and Patient 10 (neurodegenerative movement disorder phenotype with homozygous p.Leu759Alafs*4 located at the C-terminus). Immunoblotting analysis showed a significant reduction in 77-kDa PNPLA8 expression in the skin fibroblasts from Patient 1, indicating a null effect of the genetic variants (Supplemental Figure 1C). In contrast, in the case of Patient 10, the skin fibroblasts expressed the 77-kDa PNPLA8 protein, albeit at decreased levels compared to the control samples (Supplemental Figure 1D). The expression level of *PNPLA8* mRNA was not decreased in Patient 10 compared to the control samples (Supplemental Figure 1E). These results suggest that protein expression levels vary depending on the variant type, presumably leading to the phenotypic difference. Given the localization of PNPLA8 on mitochondria (10), we analyzed the acyl-chain composition profiles of mitochondrial-specific phospholipids, such as cardiolipin (CL) and monolysocardiolipin (MLCL) (29), using skin fibroblasts from Patient 10. Our investigation detected changes in the MLCL profiles of skin fibroblasts from Patient 10 (Supplemental Figure 1, F and G). Our findings suggest that despite not resulting in a null effect, the homozygous p.Leu759Alafs*4 variant in Patient 10 was associated with impaired phospholipid regulation.

Surprisingly, we identified two additional individuals from two (removed due to medRxiv policy) families with biallelic variants in *PNPLA8* from another non-neurological cohort (Families 11 and 12; Supplemental Figure 2, A and B, Supplemental Table 1). A patient from one family showed optic atrophy, and the other patient showed non-syndromic hearing loss. These patients were homozygous for distinct N-terminal frameshift variants, of which the allele frequencies were extremely low (24, 25, 27). The variants were located in the N-terminus, affecting only the largest protein isoform (MW 88 kDa) due to the multiple translation start sites in PNPLA8 (9) (Figure 2A). It is worth noting that in the family showing optic atrophy, there was an affected sibling of the proband while he was heterozygous for the variant. Although the pathogenicity of these N-terminal frameshift variants remained to be determined, our data suggest that biallelic LoF variants affecting two protein isoforms (MWs 77 and 74 kDa) are sufficient for the DEDE phenotype.

### Loss of PNPLA8 reduces the size of the subventricular zone-like region (SVZ) and the number of upper-layer neurons in cerebral organoids

From the genotype-phenotype analysis, we hypothesized that disrupted PNPLA8 function, due to biallelic truncating variants affecting the 77 and 74 kDa isoforms, lead to the DEDE phenotype, of which developmental encephalopathy and congenital microcephaly are pronounced. To test whether and how *PNPLA8* disruption affects the developing brain, we generated cerebral organoids using human induced pluripotent stem cells (iPSCs). We generated isogenic iPSC lines using sgRNAs, which were designed to target the first two exons of *PNPLA8* (Figure 3A). Using CRISPR/Cas9 genome-editing technology, the designed sgRNA and Cas9 protein were introduced into the previously established iPSC line (30–32). We obtained two sets of iPSC clones with homozygous truncating variants in *PNPLA8* (Figure 3B). We confirmed the deletion of the 77 kDa PNPLA8 protein by immunoblotting (Figure 3C). We next generated cerebral organoids from *PNPLA8* KO iPSCs and control (WT) iPSCs (33–36) (Figure 3D). The cerebral organoids continued to grow for up to 12 weeks with the appearance of multiple ventricle-like structures (Figure 3E). The organization of cortical structures, namely SOX2^+^ ventricular zone-like regions (VZ) and TUJ1^+^ neuronal areas around the VZ were confirmed (33, 37) (Supplemental Figure 3A).

**Figure 3.**
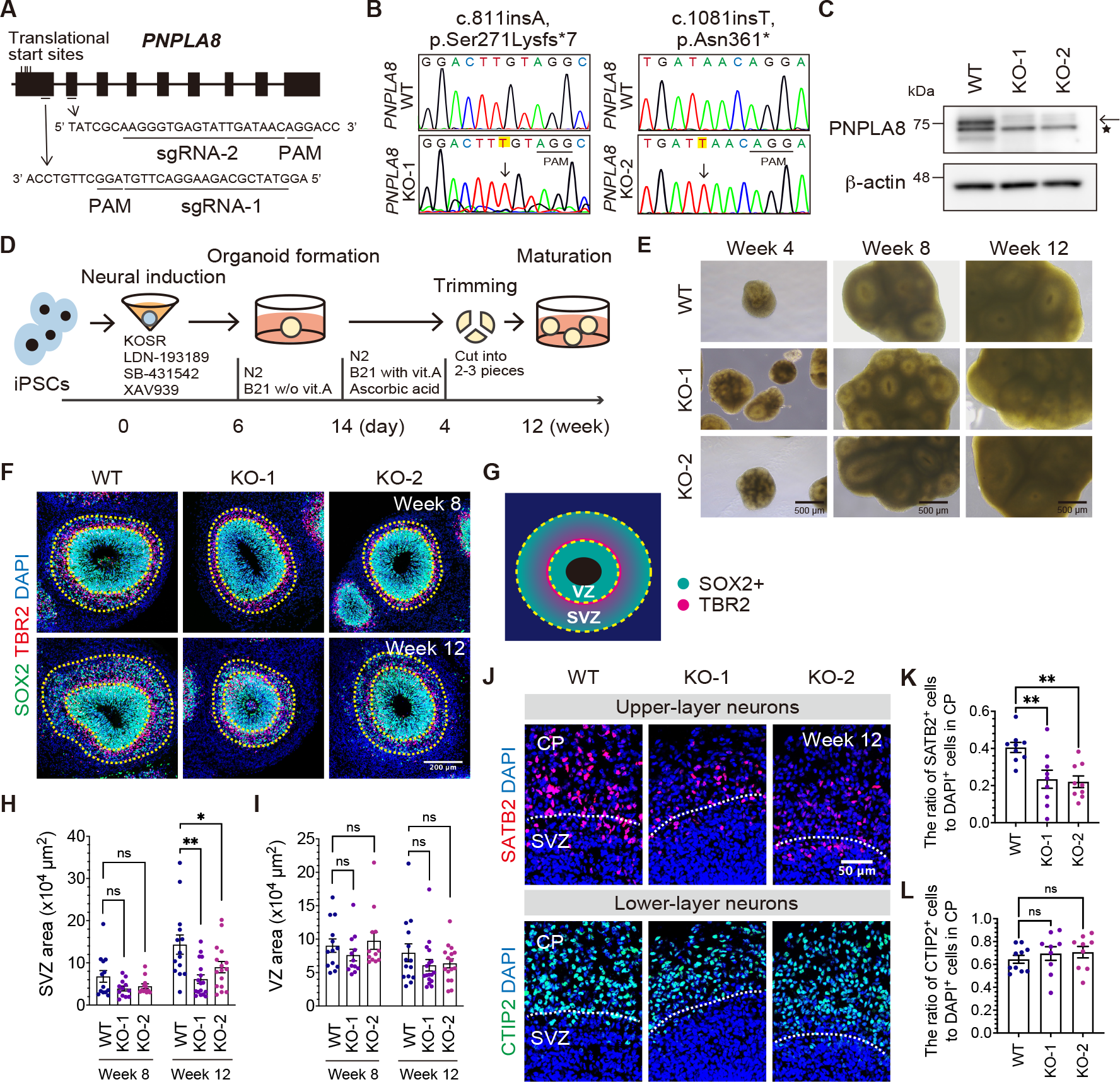
Loss of PNPLA8 reduces the size of SVZ and the number of upper-layer neurons in iPSC-derived cerebral organoids. (**A**) Schematic representation of the designed sequence of sgRNA next to protospacer adjacent motif (PAM) for *PNPLA8*. (**B**) Sanger sequencing of the CRISPR/Cas9-mediated homozygous nucleotide insertion in iPSC lines. Altered sequences are highlighted in yellow. (**C**) Immunoblotting analysis of PNPLA8 using WT and *PNPLA8* KO iPSC lines. β-actin was used as an internal protein loading control. The arrow indicates the 77 kDa PNPLA8 band, and the asterisk indicates a non-specific band. Full- length blots are shown in Supplemental Figure 8C. (**D**) Schematic illustration of the generation of cerebral organoids from *PNPLA8* KO and WT iPSC lines. KOSR and B21 indicate knockout serum replacement and Brew 21, respectively. (**E**) Bright-field microscopy images of cerebral organoids at different developmental time points. Scale bars, 500 μm. (**F** and **G**) Representative immunofluorescence images of NPC marker, SOX2, bIP marker, TBR2, and nuclear marker, DAPI at eight and 12 weeks of culture. VZ and SVZ are highlighted according to the spatial distribution of NPCs. Scale bars, 200 μm. (**H** and **I**) Quantification of the surface area of the VZ (**H**) and SVZ (**I**) at eight and 12 weeks of culture. Average values ± SEM from the number of organoids in three independent experiments (at least three organoids per experiment) are plotted. WT at week 8 (*n* = 13); WT at week 12 (*n* = 14); KO-1 at week 8 (*n* = 12); KO-1 at week 12 (*n* = 17); KO-2 at week 8 (*n* = 12); KO-2 at week 12 (*n* = 16). \**P* < 0.05, \*\**P* < 0.01; ns, not significant. One-way ANOVA followed by Dunnett’s multiple comparisons test for each time point. (**J**) Representative immunofluorescence images of CTIP2^+^ deep-layer neurons and SATB2^+^ upper-layer neurons at 12 weeks of culture. Scale bar, 50 μm. (**K** and **L**) Quantification of the CTIP2^+^ (**K**) and SATB2^+^ cells (**L**) in the cortical plate-like region (CP). Average values ± SEM from three independent experiments (at least three organoids per experiment) are plotted. WT (*n* = 9); KO-1 (*n* = 9); KO-2 (*n* = 9). \*\**P* < 0.01; ns, not significant. One-way ANOVA followed by Dunnett’s multiple comparisons test.

There was no difference in the organoid surface area between *PNPLA8* KO and WT cerebral organoids at four weeks of culture (Supplemental Figure 3B). Given that the organoid surface area was not quantifiable after four weeks of culture due to the cutting procedure (detailed in the Methods section), we next evaluated the expanding potential of the proliferative zones in the cerebral organoids. According to established terminology (38), two proliferative zones were determined; the VZ composed of densely packed SOX2^+^ aRGCs (33, 34, 37), and the SVZ, composed of sparsely distributed SOX2^+^ NPCs and TBR2^+^ bIPs (33, 34, 37) (Figure 3, F and G). We found that the SVZ area was smaller in *PNPLA8* KO cerebral organoids compared to WT cerebral organoids at 12 weeks, while there was no significant difference at eight weeks of culture (Figure 3, F and H). The size of the VZ area was similar between WT and *PNPLA8* KO cerebral organoids at both time points (Figure 3, F and I). The perimeter and thickness of the SVZ, but not the VZ, were also smaller in *PNPLA8* KO cerebral organoids than WT cerebral organoids (Figure 3F, Supplemental Figure 3, C-G). These data suggest that loss of PNPLA8 impairs the expansion of SVZ, but not VZ.

To investigate the neurogenic consequences of the reduced SVZ size in *PNPLA8* KO cerebral organoids, we quantified the number of neurons in the cortical plate-like region containing deep- and upper-layer neurons (37). The number of SATB2^+^ upper-layer neurons was significantly reduced in *PNPLA8* KO cerebral organoids compared with WT cerebral organoids (Figure 3, J and K). In contrast, there was no significant difference in the number of CTIP2^+^ deep-layer neurons (Figure 3, J and L). The decreased number of upper-layer neurons is in agreement with our findings of the reduced SVZ size in *PNPLA8* KO cerebral organoids and suggests a preferential reduction in the number of bRGCs in the SVZ, as bRGCs have been linked to the production of upper-layer neurons (2, 39, 40),

### Loss of PNPLA8 reduces the abundance of bRGCs in the SVZ

To examine which cell types are involved in the reduced expansion of the SVZ in *PNPLA8* KO cerebral organoids, we quantified the number of bRGCs and bIPs in the SVZ at 12 weeks of culture. The number of PAX6^+^HOPX^+^ bRGCs in the SVZ were significantly reduced in *PNPLA8* KO cerebral organoids compared with WT cerebral organoids (Figure 4, A and B). In contrast, the number of TBR2^+^ bIPs was similar between WT and *PNPLA8* KO cerebral organoids (Figure 4, C and D). These data suggest that the reduced abundance of bRGCs mainly contributes to the reduced size of SVZ in *PNPLA8* KO cerebral organoids.

**Figure 4.**
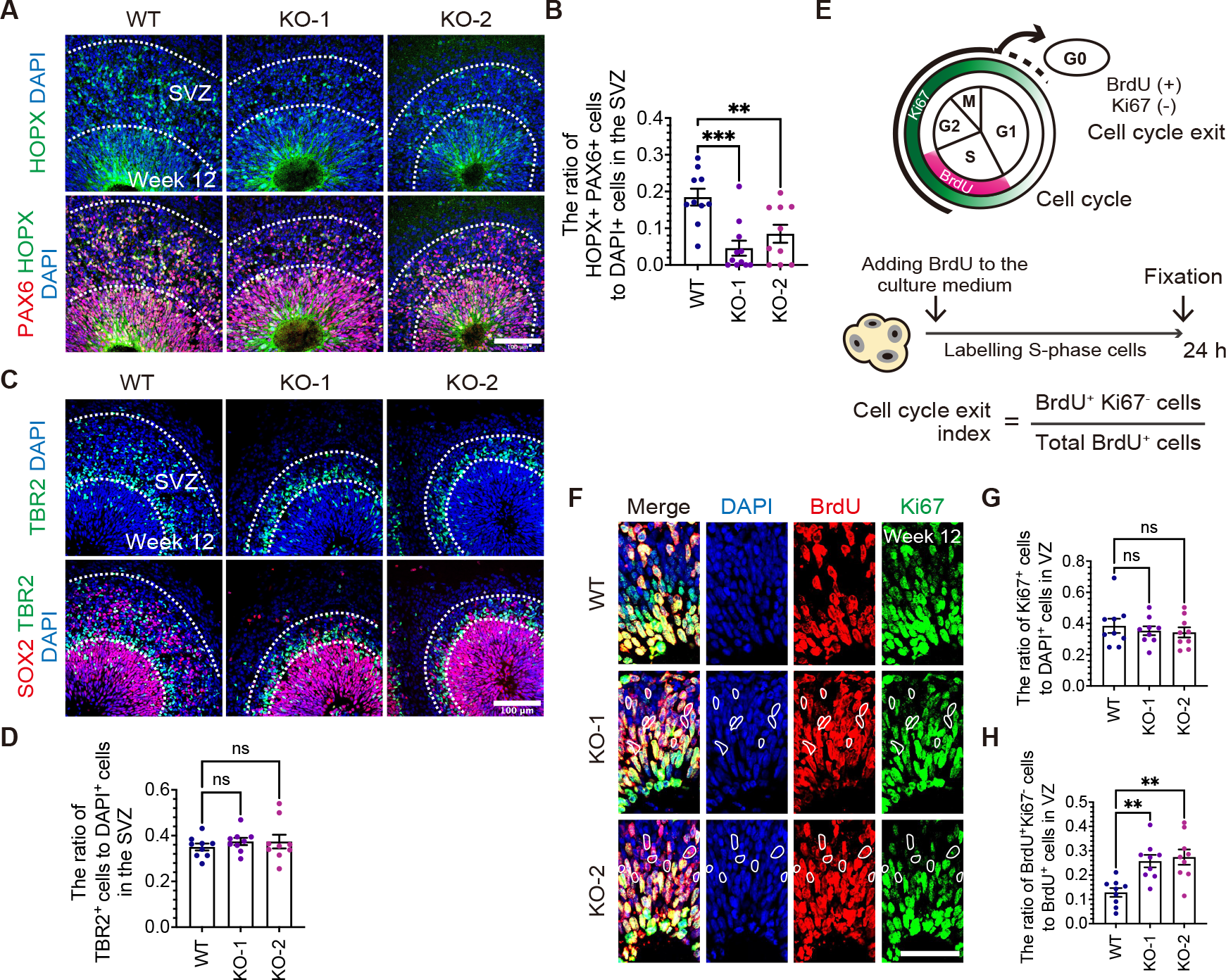
A reduced number of bRGCs and an enhanced cell cycle exit of aRGCs in *PNPLA8* KO cerebral organoids. (**A**) Representative immunofluorescence images of PAX6^+^ HOPX^+^ bRGCs at 12 weeks of culture. Scale bar, 100 μm. (**B**) Quantification of PAX6^+^ HOPX^+^bRGCs in a 100 μm-wide field of SVZ. SVZ is highlighted according to the spatial distribution of NPCs. Average values ± SEM from three independent experiments (at least three organoids per experiment) are plotted. WT (*n* = 10); KO-1 (*n* = 11); KO-2 (*n* = 10). \**P* < 0.05, \*\**P* < 0.01, \*\*\**P* < 0.001. One-way ANOVA followed by Dunnett’s multiple comparisons test. (**C**) Representative immunofluorescence images of SOX2^+^ NPCs and TBR2^+^ bIPs at 12 weeks of culture. Scale bar, 100 μm. (**D**) Quantification of TBR2^+^ bIPs in a 100 μm-wide field of SVZ. SVZ is highlighted according to the spatial distribution of NPCs. Average values ± SEM from three independent experiments (three organoids per experiment) are plotted. WT (*n* = 9); KO- 1 (*n* = 9); KO-2 (*n* = 9). ns, not significant. One-way ANOVA followed by Dunnett’s multiple comparisons test. (**E**) Schematic illustrations of the labeling paradigms for the estimation of cell cycle exit. The intensity of the green color corresponds to the abundance of the marker at a given phase, with the white color indicating an absence of the marker. Cerebral organoids were treated with BrdU to label S-phase for 24 h, and fixed for staining. (**F**) Representative immunofluorescence images of BrdU^+^ cells and Ki67^+^ cells at 12 weeks of culture. Scale bar, 50 μm. (**G** and **H**) Quantification of Ki67^+^ cells (**G**) and BrdU^+^ Ki67^−^ cells relative to BrdU^+^ cells (**H**) in the VZ. Average values ± SEM from three independent experiments (three organoids per experiment) are plotted. WT (*n* = 9); KO-1 (*n* = 9); KO-2 (*n* = 9). \*\**P* < 0.01; ns, not significant. One-way ANOVA followed by Dunnett’s multiple comparisons test.

We hypothesized that the reduced number of bRGCs could result from three possible mechanisms; the reduced proliferation of bRGCs, the increased apoptosis of bRGCs, or the decreased generation of bRGCs (41, 42). First, we quantified the number of mitotic cells in the cerebral organoids at 12 weeks of culture. The number of phospho-H3^+^ mitotic cells at the apical surface, where aRGCs divide (43), was similar between *PNPLA8* KO cerebral organoids and WT cerebral organoids (Supplemental Figure 4, A and B). The number of phospho-H3^+^SOX2^+^ NPCs in the SVZ was also similar between *PNPLA8* KO cerebral organoids and WT cerebral organoids, suggesting that the mitotic frequency was unaffected (Supplemental Figure 4, A and C). We next quantified the number of apoptotic cells in the cerebral organoids at 12 weeks of culture. The number of cleaved Caspase 3^+^ apoptotic cells in the VZ and SVZ was similar between *PNPLA8* KO cerebral organoids and WT cerebral organoids, suggesting that apoptosis was also unaffected (Supplemental Figure 4, D-F). We next analyzed the differentiation of aRGCs by cell cycle exit analysis in cerebral organoids at 12 weeks of culture. We labeled S-phase cells by a 24-h BrdU pulse and then co-immunolabelled with Ki67 and BrdU to determine cells that were exiting the cell cycle (44) (Figure 4E). There was no significant difference in the number of Ki67^+^ proliferative aRGCs in the VZ of *PNPLA8* KO cerebral organoids and WT cerebral organoids (Figure 4, F and G). The cell cycle exit index (BrdU^+^ Ki67^−^/total BrdU^+^ aRGCs) was significantly increased in *PNPLA8* KO cerebral organoids compared with WT cerebral organoids, indicative of a premature shift towards differentiating divisions of aRGCs (Figure 4, F and H). These data suggest that the altered fate specification of aRGCs might contribute to the decreased number of bRGCs in *PNPLA8* KO cerebral organoids.

### Patient-derived cerebral organoids phenocopy PNPLA8 KO cerebral organoids

Analysis of *PNPLA8* KO cerebral organoids suggested a critical role for *PNPLA8* in human brain development. However, it remained to be determined whether the *PNPLA8* variants found in patients with DEDE phenotype have the same impact on neurogenesis. We next generated iPSCs by reprogramming peripheral blood monocytes derived from Patient 1 and his parent (Control) as Patient 1 skin fibroblasts showed a significant reduction in PNPLA8 protein expression (Figure 5A). Typical iPSC morphology and capacity to differentiate into three germ layers *in vitro* were confirmed for quality assurance (45) (Supplemental Figure 5, A and B). These iPSCs carried the same variants in *PNPLA8* as the patient from which they were derived (Supplemental Figure 5C). Immunoblotting showed a significant reduction of PNPLA8 protein in Patient 1-derived iPSCs as well as *PNPLA8* KO iPSCs (Supplemental Figure 5D). The patient-derived iPSCs were induced into cerebral organoids with a slightly different protocol from that for *PNPLA8* KO cerebral organoids by using Matrigel for more efficient induction (Supplemental Figure 5E) (33, 46, 47). The cerebral organoids were cultured for eight weeks before being evaluated. Patient 1-derived cerebral organoids showed a reduced SVZ size and a slightly increased VZ size compared with Control cerebral organoids (Figure 5B, Supplemental Figure 5, F-H).

**Figure 5.**
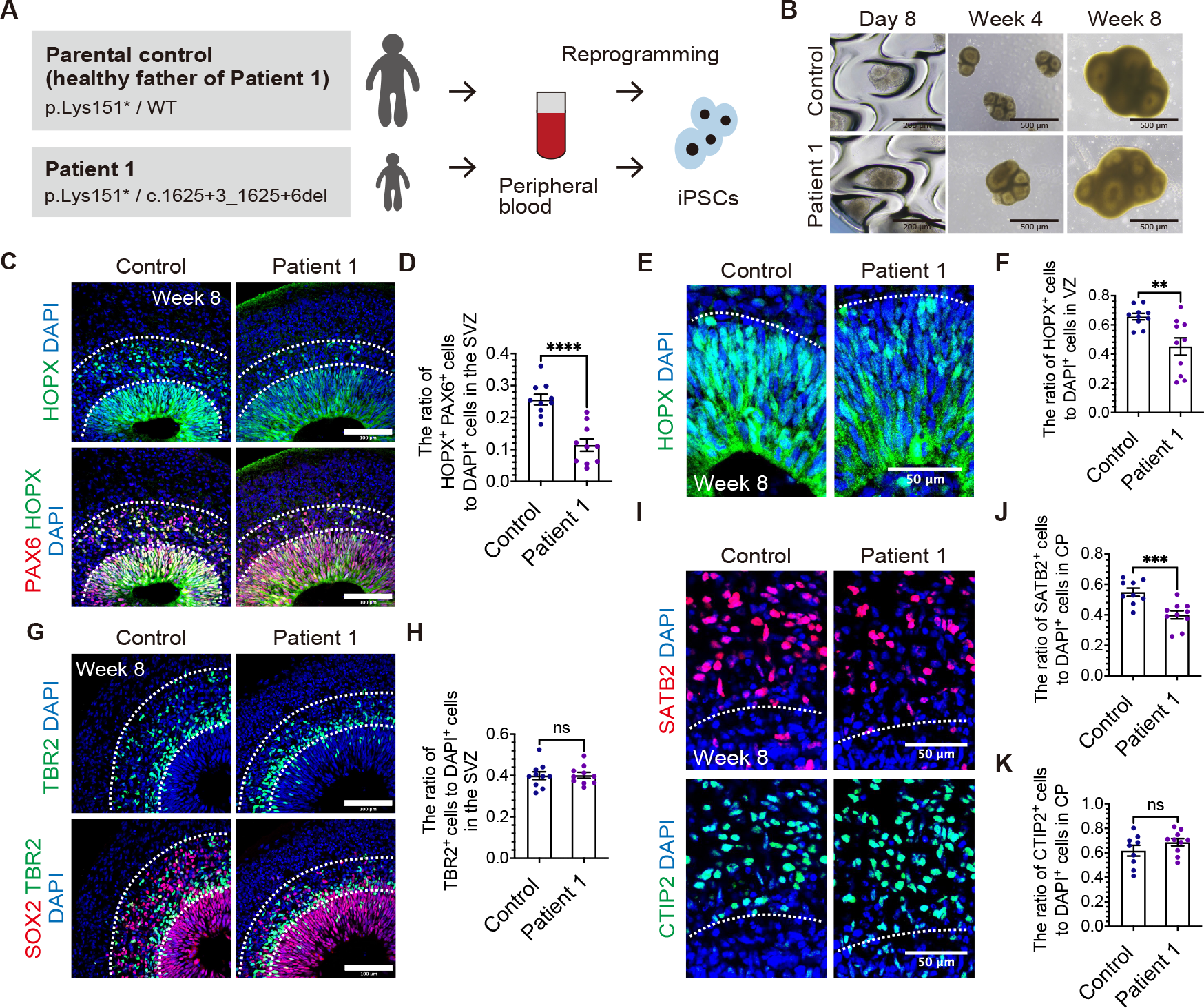
A reduced number of bRGCs and upper-layer neurons in patient iPSCs-derived cerebral organoids. (**A**) Schematic view of generating iPSCs from peripheral blood from Patient 1 and parental control. (**B**) Bright-field microscopy images of cerebral organoids at different developmental time points. Scale bars, 500 μm. (**C** and **G**) Representative immunofluorescence images of bRGCs (**C**) and bIPs (**G**) at eight weeks of culture. Scale bars, 100 μm. (**D** and **H**) Quantification of bRGCs (**D**) and bIPs (**H**) in a 100 μm-wide field of SVZ. SVZ is highlighted according to the spatial distribution of NPCs. Average values ± SEM from three independent experiments (at least three organoids per experiment) are plotted. bRGCs of Patient 1 (*n* = 10); bRGCs of control (*n* = 10); bIPs of Patient-1 (*n* = 10); bIPs of control (*n* = 10). \*\*\*\**P* < 0.0001; ns, not significant. Unpaired Student’s *t* test with Welch’s correction. (**E**) Representative immunofluorescence images of HOPX^+^ cells in the VZ at eight weeks of culture. Scale bar, 50 µm. (**F**) Quantification of HOPX^+^ cells in the VZ. Average values ± SEM from three independent experiments (at least three organoids per experiment) are plotted. Control (*n* = 10); Patient 1 (*n* = 10); \*\**P* < 0.01. Unpaired Student’s *t* test with Welch’s correction. (**I**) Representative immunofluorescence images of CTIP2^+^ deep-layer neurons and SATB2^+^ upper- layer neurons at eight weeks of culture. Scale bars, 50 μm. (**J** and **K**) Quantification of the proportion of CTIP2^+^ (**J**) and SATB2^+^ (**K**) cells in the cortical plate-like region (CP). Average values ± SEM from three independent experiments (at least three organoids per experiment) are plotted. Control (*n* = 9); Patient-1 (*n* = 10); \*\*\**P* < 0.001; ns, not significant. Unpaired Student’s *t* test with Welch’s correction.

Patient 1-derived cerebral organoids exhibited a reduced number of PAX6^+^HOPX^+^ bRGCs in the SVZ compared with Control cerebral organoids at eight weeks of culture (Figure 5, C and D). We additionally found that the number of HOPX^+^ cells in the VZ was also reduced in Patient 1-derived cerebral organoids (Figure 5, E and F). In contrast, the number of TBR2^+^ bIPs in the SVZ was similar between Patient 1-derived and Control cerebral organoids at eight weeks of culture (Figure 5, G and H). The number of SATB2^+^ upper-layer neurons was significantly reduced in Patient 1-derived cerebral organoids compared with Control cerebral organoids, while there was no significant difference in the number of CTIP2^+^ deep-layer neurons (Figure 5, I-K). These results confirmed not only the LoF nature of the *PNPLA8* variants identified in the patient, but also the phenotypic reproducibility of the cerebral organoid models derived from multiple iPS cell lines.

### Loss of PNPLA8 impairs mitochondrial function in skin fibroblasts but not in NPCs

In previous studies on mice and human patients lacking PNPLA8 function, impairment of mitochondrial function and morphology was observed in affected organs (13, 15, 22). To test whether loss of PNPLA8 affects mitochondria in an affected patient, we examined mitochondrial function and morphology in skin fibroblasts from Patient 1 (Supplemental Figure 6A). Transmission electron microscopy analysis did not reveal altered mitochondrial morphology in the patient-derived skin fibroblasts (Supplemental Figure 6B). However, live-cell assessment of mitochondrial oxidative phosphorylation (OXPHOS) activity using an extracellular flux analyzer revealed that the maximal respiration level was significantly lower in the patient-derived skin fibroblasts than those from healthy controls (Supplemental Figure 6, C and D).

Given that the function and morphology of mitochondria are involved in the fate specification of aRGCs (48), we hypothesized that the lower level of mitochondrial function caused the altered fate determination of PNPLA8-deficient aRGCs and the depletion of bRGCs. To test this, we next generated adherent monolayer cultures of *PNPLA8* KO NPCs from the same iPS cell lines we used for the cerebral organoid experiments and investigated mitochondrial morphology and function (Supplemental Figure 6E). The generated cells expressed molecular markers of NPCs and formed rosette-like structures typical for a monolayer culture of NPCs (49, 50) (Supplemental Figure 6F). These cells were used as an alternative *in vitro* model to aRGCs. There were no dysmorphic mitochondria in *PNPLA8* KO NPCs, similar to the patient-derived skin fibroblasts (Supplemental Figure 6G). Furthermore, live-cell assessment of cellular bioenergetics revealed that mitochondrial OXPHOS activity was also barely affected in *PNPLA8* KO NPCs (Supplemental Figure 6, H and I). These results suggest that loss of PNPLA8 barely affects the fate-determination process associated with mitochondrial bioenergetics and morphology in NPCs.

### Neurogenic gene expression is altered in aRGCs of patient-derived cerebral organoids

To further explore how corticogenesis is impaired in PNPLA8-deficient patients, we next performed spatial transcriptomics to assess spatially-resolved gene expression levels in aRGCs of cerebral organoids generated from Patient 1-derived iPSCs. We used a photo-isolation chemistry technique that allowed us to determine expression profiles specifically from photo-irradiated regions of interest (ROIs) (51, 52). According to the photo-isolation chemistry protocol, *in situ* reverse transcription was performed (Figure 6A) and immunostaining defined the VZ (Figure 6B). UV irradiation allowed for the VZ-specific amplification of cDNAs (Figure 6A). The library preparation using the obtained cDNAs provided more than 3.0 × 10^6^ reads per sample. These reads were mapped to more than 2.0 × 10^4^ reference genes by RNA-seq analysis. Thus, we successfully obtained assessable spatial gene expression data from cerebral organoids. Principal component analysis (PCA) showed distinct expression profiles between Control and Patient 1 samples (Figure 6C).

**Figure 6.**
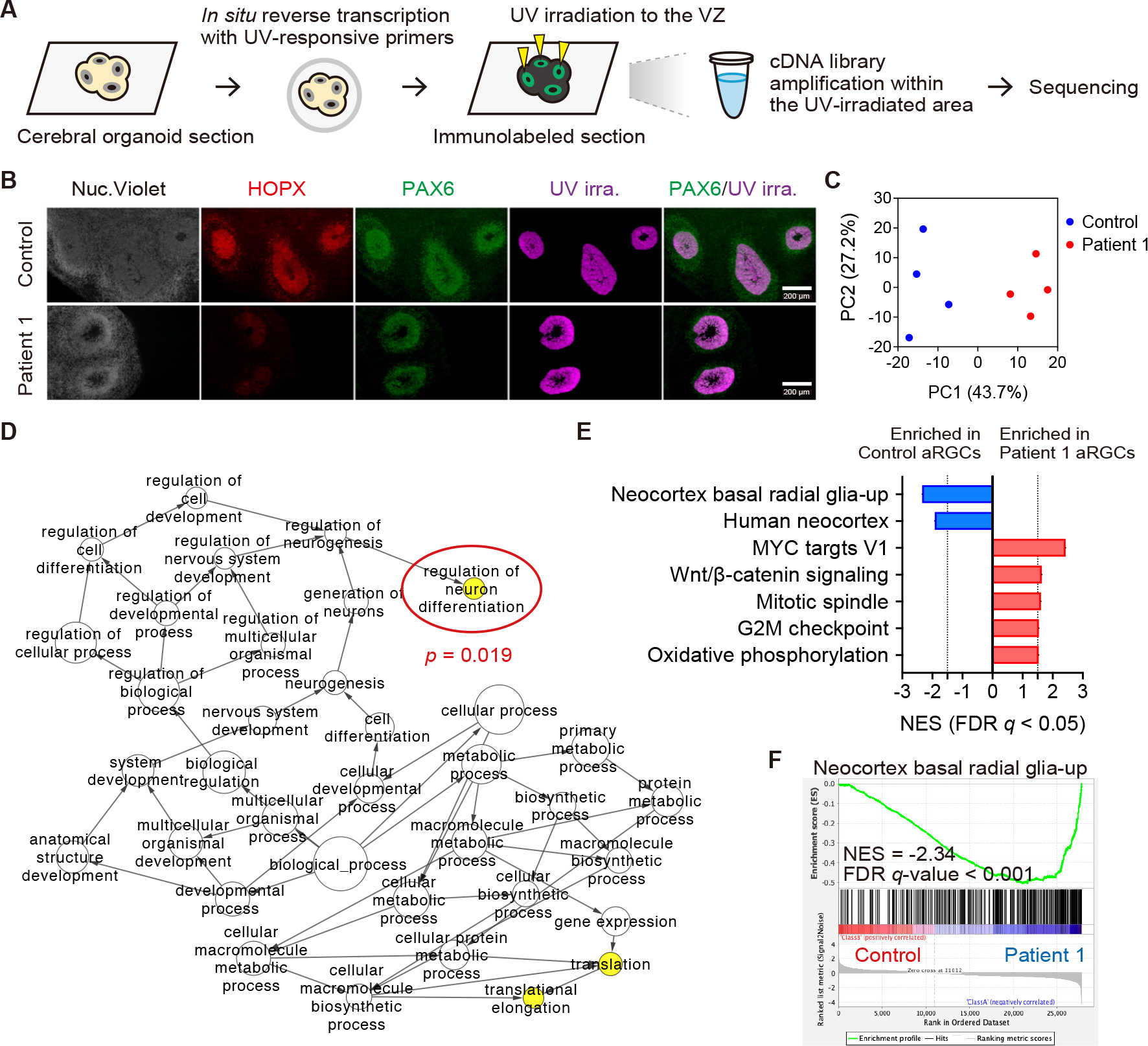
aRGCs preferentially commit to differentiated neurons instead of bRGCs in Patient 1-derived cerebral organoids. (**A**) Schematic overview of spatial transcriptomic analysis using the photo-isolation chemistry technique. (**B**) Representative immunofluorescence images of cerebral organoids at eight weeks of culture. VZ and SVZ are visualized with PAX6 and HOPX. The VZ was irradiated with UV light where PAX6^+^ RGCs were densely packed. Merged images of immunofluorescence and UV-irradiated area are shown in the right-most panels. Four distinct lines of cerebral organoids were utilized. The RNA samples were extracted from at least four VZs from at least two organoids per sample. (**C**) PCA for the expression profiles. PC1 explained 43.7% and PC2 explained 27.2% of the variation. (**D**) GO-term networks generated by BiNGO enrichment analysis for DEGs of patient aRGCs. Node size represents the GO hierarchy. The yellow color of the nodes represents significant enrichment levels with *P* < 0.05. (**E**) GSEA results ranked by normalized enrichment score (NES) using Curated gene sets and Hallmark gene sets of MSigDB signatures. Negative NES, depleted signature in Patient 1; positive NES, enriched signature in Patient 1. NES cutoff > 1.5 or < −1.5. FDR *q*-value cutoff < 0.05. (**F**) Enrichment plot for “Neocortex basal radial glia-up”, showing the profile of the running enrichment score and positions of gene set members on the rank-ordered list.

To characterize gene expression profiles, we detected differentially-expressed genes (DEGs) from the data. As shown in the heatmap of DEG clustering, there were 104 up- and 43 down-regulated genes in Patient 1, compared with the Control (Supplemental Figure 7A, Supplemental Table 2). The DEGs were analyzed using Cytoscape-based BiNGO software for enrichment analysis (53). This analysis revealed three significantly enriched biological processes, such as “translation”, “translational elongation” and “regulation of neuron differentiation” (Figure 6D). These results suggest that patient aRGCs exhibited a premature shift towards differentiating divisions. Next, we performed gene set enrichment analysis (GSEA) and determined significantly different expressions of gene sets from the mSigDB database (54–56). Among the acquired data, we found that gene sets related to “Neocortex basal radial glia-up” and “Human neocortex” were much less enriched in Patient 1-derived cerebral organoids (57) (Figure 6, E and F). These results support our notion that loss of PNPLA8 impairs bRGC-mediated neurogenesis by altering the fate specification of aRGCs. We next created an enrichment map to visualize GSEA results using the Gene Ontology (GO) tool as a gene-set source (58). This visualization confirmed the enrichment of gene sets related to neuron development (e.g., “regulation of axonogenesis”) in Patient 1-derived cerebral organoids (Supplemental Figure 7B). The enriched gene sets in Patient 1 also exhibited diverse molecular functions, including RNA splicing (e.g., “regulation of RNA splicing”), chromatin organization (e.g., “chromatin remodeling”), mitochondrial organization (e.g., “protein localization to mitochondrion”), and organelle development (e.g., “ribosome”) (Supplemental Figure 7B).

We further identified gene sets related to cell fate specification. Regarding the cell cycle, the GSEA results showed enrichment of multiple cell cycle-related gene sets, such as MYC targets, mitotic spindle, and G2/M checkpoint (Figure 6E, Supplemental Figure 7C) in Patient 1-derived cerebral organoids, suggesting that cell cycle progression was promoted by PNPLA8 deficiency (59, 60). The enrichment of WNT/β-catenin signaling in patient aRGCs suggested preferential proliferation rather than differentiation, but was similar to the specific subtype of NPCs that is directed toward differentiation into deep-layer neurons (61) (Figure 6E). Regarding metabolism, gene sets related to mitochondrial OXPHOS and mitochondrial organization were enriched in Patient 1-derived cerebral organoids, consistent with evidence that the differentiation of NPCs to a neuronal lineage is accompanied by a metabolic switch from glycolytic metabolism to mitochondrial OXPHOS (48, 62, 63) (Figure 6E, Supplemental Figure 7D).

### Loss of PNPLA8 dysregulates neurogenic signaling and phospholipid metabolism

Spatial transcriptomic analysis revealed the altered fate commitment of aRGCs in patients with *PNPLA8* variants. We next analyzed pathways involved in the neurodevelopmental defects of PNPLA8-deficient patients using the transcriptomics data. The Pathway Interaction Database, a collection of curated pathways focusing on signaling and regulatory pathways rather than metabolic processes or generic mechanisms like transcription and translation (64), revealed “lysophospholipid pathway” as the top-ranked enrichment in Patient 1-derived cerebral organoids (Figure 7, A and B). Within the dataset, *LPAR2* was significantly enriched in Patient 1, whereas *LPAR1* was not enriched in Patient 1 (Figure 7C). Lysophosphatidic acid (LPA) acts as a signaling molecule through its distinct G protein-coupled receptors (LPAR1-6) with diverse effects during brain development (65, 66). Specifically, LPAR2 is thought to be involved in neural differentiation, whereas LPAR1 is likely to mediate various responses, such as proliferation (66, 67). These results suggest the possible link between dysregulated neurogenic signaling pathways and the aberrant phospholipid metabolism in PNPLA8- deficient aRGCs.

**Figure 7.**
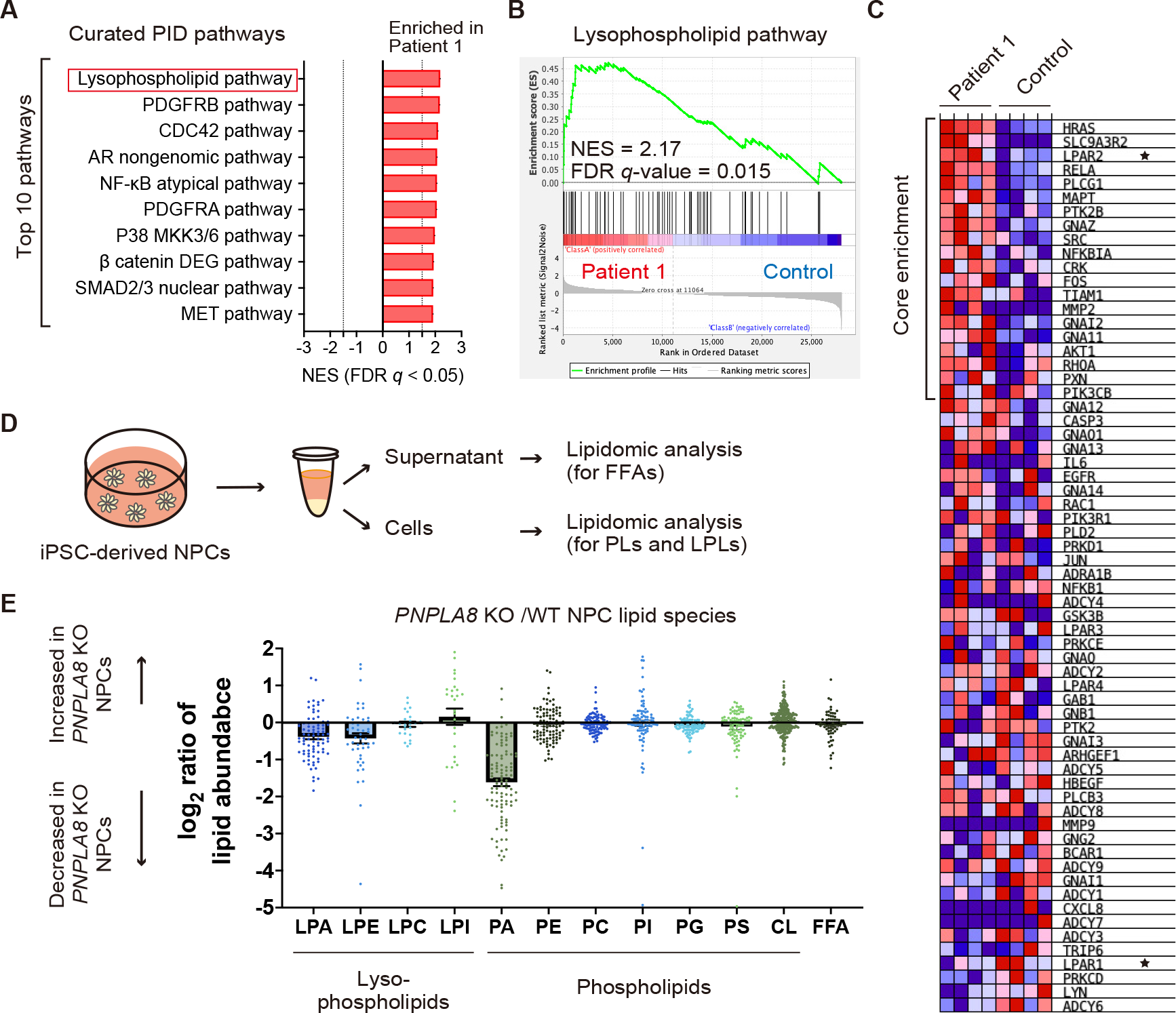
Loss of PNPLA8 affects phospholipid metabolism. (**A**) GSEA results ranked by NES using the Pathway Interaction Database gene sets of MSigDB signatures. Positive NES, enriched signature in Patient 1. NES cutoff > 1.5. FDR *q*-value cutoff < 0.05. (**B**) Enrichment plot showing “lysophospholipid pathway”. (**C**) Heat map of the marker genes from the “lysophospholipid pathway” gene set in the comparison of Patient 1 aRGCs (left columns) vs. control aRGCs (right columns). Expression values are represented as colors and range from red (highest expression), pink (moderate), light blue (low), to dark blue (lowest expression). The core enrichment genes, which contribute most to the enrichment result of the gene set, were determined by the running enrichment scores. (**D**) iPSC-derived NPCs and culture supernatants were subjected to lipidomic analysis to quantify phospholipids (PLs), lysophospholipids (LPLs), and free fatty acids (FFAs). (**E**) Individual lipid species detected in lipid extracts from WT NPCs or *PNPLA8* KO NPCs, grouped by class. The log2 average ratios of liquid chromatography-mass spectrometry/mass spectrometry profiles are shown. Data from *PNPLA8* KO-1 and KO-2 NPCs are combined as “KO”. Bars represent the mean fold enrichment for individual lipid species in that class ± SEM for four independent experiments. Each dot indicates a single measurement of LPA (*n* = 83), LPE (*n* = 47), lysophosphatidylcholine (LPC ; *n* = 24), lysophosphatidylinositol (LPI; *n* = 27), PA (*n* = 104), phosphatidylethanolamine (PE; *n* = 101), phosphatidylcholine (PC; *n* = 104), phosphatidylinositol (PI; *n* = 92), phosphatidylglycerol (PG; *n* = 104), phosphatidylserine (PS; *n* = 89), cardiolipin (CL; *n* = 280), and FFA (*n* = 64).

To examine whether loss of PNPLA8 affects phospholipid metabolism, we next performed lipidomic analysis using iPSC-derived NPCs and culture supernatants (Figure 7D). The amounts of LPA, lysophosphatidylethanolamine (LPE), and phosphatidic acid (PA) were decreased in *PNPLA8* KO NPCs compared with WT NPCs, while there was no significant difference in that of other phospholipids and FFAs (Figure 7E). These results suggest that loss of PNPLA8 reduces the synthesis of LPA, LPE, and PA through decreased PLA2 activity. The dysregulation of phospholipid metabolism, including LPA synthesis, is consistent with the possibility that PNPLA8 contributes to the fate-determination process through the membrane remodeling of NPCs.

## Discussion

In this study, we provide insight into the understanding of *PNPLA8*-related neurological disease. This insight is based on the clinical and genetic evaluation of newly identified patients with *PNPLA8* variants. Our study demonstrates that biallelic null variants in *PNPLA8* cause microcephaly through the reduced abundance of bRGCs. Our findings from the combination of iPSC-based models and multi-omics analysis suggest that PNPLA8 contributes to membrane remodeling and the fate-determination process in aRGCs.

Our data expand the spectrum of individuals with biallelic *PNPLA8* variants, which reveals a continuum ranging from a childhood-onset neurodegenerative movement disorder to developmental and epileptic-dyskinetic encephalopathy, termed DEDE. Besides the phenotypic variability, our data allowed for the detailed evaluation of genotype-phenotype correlations. The patients with childhood-onset neurodegenerative movement disorders commonly carried the homozygous recurrent C-terminal frameshift variant (c.2275_2276del; p.Leu759Alafs*4). According to the ‘nonsense-mediated mRNA decay (NMD) rule’, premature termination codons located 55 nucleotides or more downstream of the last exon-exon junction are considered to escape from NMD, allowing for the translation of truncated protein (68). Considering the position where the premature termination codon is introduced, the C-terminal truncating variant theoretically avoids NMD, presumably leading to the residual enzymatic activity (68, 69). Our study provides evidence supporting this hypothesis through the analysis of cells derived from Patient 10, which exhibited altered MLCL acyl-chain profiles with a detectable expression of PNPLA8 protein (Supplemental Figure 1, D-G). This unreported phenotype expands the mild end of the spectrum as they showed milder intellectual disability and the absence of seizures as well as their late onset at adulthood. This phenotype highlights the aspects of a neurodegenerative movement disorder in *PNPLA8*-related neurological disease.

In contrast, patients with the DEDE phenotype carried biallelic null variants affecting at least the 77 and 74 kDa protein isoforms. Considering the NMD mechanism (68) with evidence of the deletion of the 77 kDa protein in Patient 1 (Supplemental Figure 1C), complete loss of PNPLA8 function is likely to result in the severe end of the phenotypic spectrum, of which distinct features are early-onset severe neurological manifestations and congenital microcephaly with brain malformations. The three severe cases previously reported fall into this category; two girls with homozygous nonsense variants (20, 23), and a girl with homozygous missense variants, which presumably disrupts protein function due to the location of the variant in the functional domain (21). The remaining reported cases seem to fall into the intermediate category (22, 23). Consistently, they carried the combination of the following variants: one was an LoF variant, the other was the recurrent C-terminal truncating variant or missense variant with a possible mild effect (22, 23). Collectively, it is likely that the phenotypic features vary depending on the residual protein function. Further studies are needed to determine the pathogenicity of each variant, especially the novel missense variant identified in the patient from Family 8 showing an intermediate phenotype.

The identification of these patients led to our initial hypothesis that complete loss of PNPLA8 function leads to neurodevelopmental deficits, including cortical malformations. Cerebral organoids lacking PNPLA8 showed a reduced abundance of bRGCs, in contrast to bIPs. In line with this, we observed a specific decrease in the number of upper-layer neurons (70). The previous postmortem study of a 27-week-old fetus with microcephaly with simplified gyral pattern showed a proliferative defect in the outer SVZ, where bRGCs exist, and a lack of upper-layer neurons as the characteristic histological findings (71). Similar pathomechanisms were experimentally shown in *PNPLA8* KO cerebral organoids, suggesting that loss of PNPLA8 results in microcephaly with a simplified gyral pattern. Interestingly, our findings appear inconsistent with the previous reports on *Pnpla8* KO mice as they showed a normal brain morphology and no apparent neurodevelopmental phenotype (12, 13). However, this phenotypic discrepancy could be explained by the differences in bRGC abundance between humans and mice (72, 73). Our data suggest that PNPLA8 has a critical role in the bRGC- mediated cortical expansion in addition to the conserved role in the maintenance of brain function (11).

Based on the recent evidence that mitochondrial function and morphology influence the fate decisions of NPCs (48), we hypothesized that dysfunctional or dysmorphic mitochondria underlie the enhanced cell cycle exit observed in *PNPLA8* KO cerebral organoids. However, loss of PNPLA8 affected neither function nor morphology of mitochondria in NPCs, while there was a significant reduction of the maximum oxygen consumption in patient-derived skin fibroblasts. This discrepancy could be attributed to the energetic dependency since NPCs are predominantly dependent on glycolytic metabolism rather than OXPHOS (48). Given that neurons are dependent on OXPHOS (48), mitochondrial dysfunction may also occur in neurons, thereby contributing to the neurodegeneration in *PNPLA8*-related diseases. Our data indicate that loss of PNPLA8 appears to have little effect on NPC mitochondria and the fate-determining mechanisms in which mitochondria are involved.

By using a photo-isolation chemistry technique, we provide spatially resolved *in situ* transcriptional profiles of cerebral organoid sections. Our data indicate that aRGCs lacking PNPLA8 preferentially committed to differentiated neurons instead of bRGCs in cerebral organoids. Recent evidence indicates that the transcriptional state of bRGC emerges in the VZ during early cortical neurogenesis (74). Therefore, our findings suggest a reduced production of bRGCs from aRGCs based on gene expression profiles. Premature neural differentiation of the aRGCs is known as one of the major mechanisms underlying the microcephalic phenotype (75). In addition, bRGC abundance has been linked to cortical expansion and gyrification (72).

It is likely that both premature neural differentiation and reduced production of bRGCs underlie the pathogenesis of microcephaly with simplified gyral pattern in *PNPLA8*-related neurological disease. The upregulation of lysophospholipid signaling genes in patient-derived cerebral organoids suggests the contribution of PNPLA8 to the fate-decision process with its membrane remodeling capacity. It needs to be further addressed which pathway is involved in the pathogenesis through rescue experiments.

Our lipidomics data reveal the reduced abundance of LPA, LPE, and PA in *PNPLA8* KO NPCs, which supports our notion that loss of PNPLA8 impairs membrane remodeling in NPCs. In line with the unaffected morphology and function of mitochondria, the amount of CL was also unaffected in *PNPLA8* KO NPCs. The reduction of LPA and LPE can be explained by decreased PLA2 activity (76). However, the significant reduction of both LPA and PA suggests the impaired *de novo* synthesis of phospholipids (77). Given that LPE can be catabolized to glycerol 3-phosphate, which is a starting material for the *de novo* synthesis of phospholipids, the reduction of LPE might also affect PA synthesis (77). Collectively, it is likely that loss of PNPLA8 impairs phospholipid catabolism leading to a reduced synthesis of phospholipids in NPCs. Pinson *et al*. discussed that fatty acids are building blocks for membrane synthesis and are essential for bRGC proliferation in the developing modern human brain (78). It therefore appears worthwhile to investigate whether PNPLA8-dependent phospholipid synthesis may also underlie bRGC production as building blocks for membrane synthesis.

## Methods

Additional details can be found in Supplemental Methods.

### Patients

Fourteen individuals from 12 unrelated families from around the world were identified through international collaboration and data sharing. Although Family 6 and 7 in this study had been previously reported without details (20, 79), we included these patients with unreported clinical and genetic data. Additionally, we reviewed the clinical features of previously reported patients (20–23) (Table 1). Physical features of weight and head circumference were converted to standard deviation scores, corrected for gender, age, and gestational age (80, 81). Clinically-acquired brain images were available for eight patients. Among them, only low-resolution photographs of brain MRIs were available for Patient 7 and Patient 10.

### Genetic analysis

Proband-only or trio-exome/genome sequencing and bioinformatics with subsequent candidate variant Sanger segregation analysis was carried out on DNA extracted from blood-derived leukocytes at different genetic diagnostic and research laboratories worldwide following slightly different protocols as described previously (82–84). Nucleotide sequences were described with reference to *PNPLA8* transcript NM_001256007.3.

### Generation of iPSC KO cell lines

Previously established human iPSCs (Windy) were used in this study (30–32). CRISPR/Cas9-mediated genome editing in iPSCs was performed as previously described with modifications (85). For the preparation of ribonucleoprotein (RNP) complex, 2 µL of 100 µM CRISPR RNA and 2 µL of 100 µM trans-activating CRISPR RNA with Atto550 was mixed, followed by incubation at 95°C for 5 min and then at room temperature (RT) to form sgRNA complex. Then, 1 µL of 62 µM Cas9 protein was added to the gRNA complex and incubated at RT for 30 min to form an RNP complex. Subsequently, 1 × 10^6^ iPSCs were electroporated with 5 µL of the prepared RNP complex dissolved in 95 µL of Opti-MEM (NEPA21; Nepagene, Japan). Single clones of Atto550^+^ iPSCs were isolated with a cell sorter (FACS Aria II; BD Biosciences, USA). After the expansion of the clones, we extracted genomic DNA and confirmed the homozygous KO state by Sanger sequencing. Primer sequences are provided in Supplemental Table 3.

### iPSC cell culture

Human iPSCs were maintained in Stemfit with iMatrix-511 (0.2 μg/cm^2^). iMatrix-511 was used in an uncoated manner (86). Every four to seven days, the cells were passaged using accutase. The cells were seeded at 555 cells/cm^2^ density onto a culture dish containing Stemfit, iMatrix, and 10 μM of Y-27632. One day after passaging, the culture medium was replaced with Stemfit without Y-27632. The culture medium was changed every one to three days. Human iPSCs (Windy), patient iPSCs, and the control iPSCs were used between passages 40 to 60, 30 to 50, and 30 to 50, respectively.

### Patient-derived iPSC cell line generation

Patient and control-derived iPSCs were generated according to published protocols (87–90). In brief, peripheral blood mononuclear cells were collected in the same manner as with lymphoblastoid cell line generation. Cells were cultured in StemSpan H3000 supplemented with recombinant human interleukin-6 (0.1 µg/mL), recombinant human stem cell factor (0.3 µg/mL), recombinant human thrombopoietin (0.3 µg/mL), recombinant human Flt3 ligand (0.3 µg/mL), and recombinant human interleukin-3 (0.01 µg/mL). Cells were transfected by electroporation (Nucleofector-2b; Lonza, Germany) using a Human CD34+ Cell Nucleofector Kit with 0.63 μg of each episomal vector (pCE- hOCT3/4, pCE-hSK, pCE-hUL, pCE-mp53DD, and pCXB-EBNA1) (90). Following transfection, cells were cultured on mitomycin C-treated mouse embryonic fibroblast feeder cells in iPSC medium consisting of DMEM/F12 supplemented with 20% Knockout Serum Replacement, 2 mM L-glutamine, 0.1 mM nonessential amino acids, 0.1 mM 2- mercaptoethanol, 100 units/mL penicillin, and 100 μg/mL streptomycin, and 4 ng/ml FGF-2. Mouse embryonic fibroblasts were prepared from embryonic day 13-14 of Slc:ICR mouse embryos. Differentiation potentials of iPSCs were determined as previously described (91).

Briefly, for embryoid body (EB) formation, dissociated iPSCs were seeded and cultured in DMEM/F12 containing 5% knockout serum replacement, 2 mM L-glutamine, 0.1 mM MEM non-essential amino acids, 0.1 mM 2-ME, 10 μM Y-27632, 100 U/mL penicillin, and 100 μg/mL streptomycin. After seven days of floating culture, the EBs were transferred onto gelatin-coated plates and cultured in DMEM containing 10% FBS for another seven days to induce spontaneous differentiation. After fixation with 4% paraformaldehyde in PBS for 15 min, the expression of differentiation markers was confirmed by immunocytochemistry. The absence of pathogenic chromosomal copy number variation was confirmed by microarray-based comparative genomic hybridization analysis as previously described (87).

### Generation of cerebral organoids from iPSCs

To generate *PNPLA8* KO cerebral organoids, we used a dual SMAD inhibitor protocol with slight modifications (35, 36). We seeded 3,000 iPSCs into 96-well V-bottom plates and cultured them in the following ‘induction media’ for the first six days with replacement every two days: DMEM/F12 containing 20% knockout serum replacement, 1 mM MEM non-essential amino acids, 2 mM L-glutamine, 100 µM 2-ME, 100 nM LDN-193189, 10 µM SB-431542, 2 µM XAV939, and 50 µM Y-27632 (for the first 48 h). On the sixth day in suspension, the spheroids were transferred into non-adherent 24-well plates, pre-coated with poly 2-hydroxyethyl methacrylate solution (1.2 g of 2-hydroxyethyl methacrylate in 50 mL of 95% ethanol). The spheroids were then cultured in the following ‘organoid culture media 1’: 50% DMEM/F12, 50% MACS Neuro Medium, 1 mM MEM non-essential amino acids, 2 mM L-glutamine, 50 µM 2-ME, 400 nM insulin, 0.5× N-2 MAX Media Supplement, 100 U/mL penicillin, 100 μg/mL streptomycin, and 0.5× MACS Brew21 minus Vitamin A replaced every two to three days. The plates were rotated on an orbital shaker (CS-LR; TAITEC, Japan) at 70 rpm. To promote differentiation of the neural progenitors into neurons, the spheroids were cultured in ‘organoid culture media 2’: 50% DMEM/F12, 50% MACS Neuro Medium, 1 mM MEM non-essential amino acids, 2 mM L-glutamine, 50 µM 2-ME, 400 nM insulin, 0.5× N2 supplement, 100 U/mL penicillin, 100 μg/mL streptomycin, 0.5× MACS Brew21 with Vitamin A, and 200 µM L-ascorbic acid from day 14 up to three months. The culture media was replaced every one to three days. After one month, the organoids were cut into two to three pieces with a scalpel under a stereo microscope (S6E; Leica, Germany) every week to prevent cell death in the central portions (92).

Patient and control-derived cerebral organoids were generated by a slightly different protocol. For EB formation, we seeded 188,000 iPSCs into EZ-sphere 24-well plates (about 400 cells per micro well) and cultured them in the ‘EB formation media’: DMEM/F12 containing 20% knockout serum replacement, 1 mM MEM non-essential amino acids, 2 mM L-glutamine, 4 ng/mL FGF-2 (93), and 50 µM Y-27632 (for the first 48 h). On the third day, the culture media was changed to ‘induction media’ until the sixth day, with replacement every two days. On the sixth day, the culture media was removed, and EB spheroids were embedded in 200 µL of cold Matrigel and incubated at 37°C for 30 min. Subsequently, 1 mL of ‘organoid culture media 1’ was added to the culture plates. On the 10th day, the cerebral organoids were transferred into six-well 2-hydroxyethyl methacrylate-coated plates on an orbital shaker at 70 rpm. On the 14th day, culture media was replaced with ‘organoid culture media 2’. The cerebral organoids were cultured for up to two months. The culture media was replaced every one to three days. After one month, the organoids were cut into two to three pieces with a scalpel under a stereo microscope every week to prevent cell death in the central portions (92).

Images of the organoids were acquired using an inverted optical microscope (CKS53; Olympus, Japan). The organoid size was measured based on the surface of the organoids using ImageJ and FIJI software.

### Generation of iPSC-derived NPCs

Generation of NPCs from iPSCs was performed as previously described with modifications (44, 94). Briefly, 188,000 cells were seeded into EZ- sphere 24-well plates (about 400 cells per micro well) and cultured in ‘induction media’ for the first six days with replacement every two days. For the first 48 h, 50 µM Y-27632 was added. On the sixth day, spheroids were seeded onto six-well plates that had been coated with poly-L-lysine for 1 h and subsequently with laminin overnight. The attached spheroids were cultured in the following ‘neural media’: 50% DMEM/F12 and 50% MACS Neuro Medium containing 1 mM MEM non-essential amino acids, 2 mM L-glutamine, 0.5× N-2 MAX media supplement, 0.5× MACS Brew21, 20 ng/mL FGF-2, 20 ng/mL EGF, 100 U/mL penicillin, and 100 μg/mL streptomycin. The spheroid-derived monolayer cells were cultured until they formed neural rosettes, with the replacement of culture media every two days. Subsequently, whole cells were dissociated, passaged into a new dish, and/or cryopreserved in Stem Cell Banker. The thawed cells were seeded onto poly-L-lysine and laminin-coated plates filled with ‘neural media’ and confirmed to be able to form neural rosettes. iPSC-derived NPCs were passaged once a week and used for each experiment on passages three to five. To analyze NPCs for immunocytochemistry, iPSC-derived NPCs grown on a 96-well glass bottom plate were fixed with 4% paraformaldehyde in PBS for 15 min.

### Immunofluorescence

For cryosections, cerebral organoids were fixed in 4% paraformaldehyde in PBS at 4°C overnight, followed by 30% sucrose in PBS at 4°C for 24 h. Next, the organoids were transferred into Tissue-Tek O.C.T. compound and frozen at −80°C. Sixteen μm-thick sections were obtained using a cryostat (CM3050; Leica, Germany). Cryosections were incubated in PBS to wash away the OCT. Cryosections and cell monolayers were permeabilized in 5% Triton X-100 in PBS at RT for 5 min, and blocked in Blocking-One at RT for 1 h. Sections or cells were incubated at 4°C overnight with primary antibodies diluted in Blocking-One. After three PBS washes, samples were incubated in secondary antibodies and DAPI, which were diluted in Blocking-One. After PBS washes, samples were mounted with Fluoromount-G. For BrdU staining, sections were treated with 2N HCl at 37°C for 20 min, followed by three PBS washes before permeabilization. Images were obtained with a SpinSR10 confocal microscope and cellSens software (Olympus, Japan), and an A1RS^+^ confocal microscope and NIS Elements software (NIKON, Japan) with 10×, 20×, or 40× magnification. Quantification of images was performed using FIJI software. Primary and secondary antibodies are listed in SupplementalTable 3.

### Spatial transcriptomic analysis

Cerebral organoids were fixed in 4% paraformaldehyde in PBS at RT for 10 min, followed by 30% sucrose in PBS at 4°C for 24 h. The organoids were transferred into Tissue-Tek O.C.T. compound, rapidly frozen in dry ice-cooled 2-methylbutane, and kept frozen at −80°C. Subsequently, 10 μm-thick sections were obtained using a cryostat. Spatial transcriptomic analysis using the photo-isolation chemistry technique was performed as previously described (51, 52). Briefly, reverse transcription was performed by adding UV-responsive 6-nitropiperonyloxymethyl-caged reverse transcription primers containing T7 promotor, unique molecular identifiers (UMIs), multiple barcodes, and poly T sequence onto the organoid sections. Immunofluorescence was then performed to visualize ROIs for subsequent UV irradiation. To cleave 6-nitropiperonyloxymethyl moieties from reverse transcription primers, the ROIs were irradiated with UV light for 15 min with a Digital Micromirror Device (Polygon 1000-G; Mightex Systems, Japan). The total tissue lysate was then collected and purified using 20 mg/mL proteinase K. Second-strand DNA was synthesized by the nick translation method. For *in vitro* transcription reaction, synthesized cDNAs were transcribed to RNAs using a T7 Transcription Kit. After the collection of ROI-specific RNAs, the RNAs were further reverse-transcribed, followed by paired-end sequencing on the Illumina platform (Read 1: UMIs and barcode, Read 2: cDNA). Sequences were separated by the sample barcodes with UMI-tools and mapped to the reference genome using HISAT2. UMI-tools and featureCounts were used to generate UMI count data assigned to genes. DEGs were extracted by DESeq2 (FDR = 0.1). The DEGs were further filtered with adjusted *P*-values (padj) < 0.1. DESeq2 was also used to transform the count data into regularized log data before performing

PCA using the R prcomp function. Enrichment maps were constructed with Cytoscape 3.9 and the BiNGO plug-in using the default settings (53). The degree of enrichment for each GO was assessed and considered significant when *P* < 0.05. GSEA was performed on normalized counts of RNA-seq datasets. Enrichment *P*-values were estimated by 1,000 permutations. The gene set databases used were Curated (C2; *n* = 6,449), Hallmark (H; *n* = 50), Gene Ontology (C5; *n* = 15,703), and Pathway Interaction Database (C2; PID; *n* = 196) gene set collections (54, 55, 57, 64). Specifically, GO results were visualized using the Enrichment Map plug-in for Cytoscape 3.9 (*P*-value < 0.005, FDR *q*-value < 0.05). Clusters of functionally related enriched GO terms with ≥ 5 nodes were manually detected.

### Lipidomic analysis

Lipidomic analysis was performed by electrospray ionization-liquid chromatography-mass spectrometry/mass spectrometry according to a published protocol (95). Lipids from each sample were extracted by the method of Bligh and Dyer (96). To determine the amount of each phospholipid, lipid phosphorus was measured by Bartlett’s method (97). The analysis of phospholipid species was performed using a hybrid triple quadrupole-linear ion trap mass spectrometer (4500Q-TRAP; AB Sciex, Japan) with Nexera UPLC system (Shimazu, Japan). For internal standards, the phospholipid mixture containing 12.5 pmol of each phospholipid (phosphatidylcholine 25:0, phosphatidylethanolamine 25:0, phosphatidylglycerol 25:0, phosphatidylinositol 25:0, phosphatidylserine 25:0, and PA 34:0), 25 pmol of each lysophospholipid (lysophosphatidylcholine-d49 16:0, LPE-d7 18:1, lysophosphatidylglycerol 17:1, and lysophosphatidylserine 17:1), 250 pmol of lysophosphatidylinositol 17:1, 75 pmol of LPA 17:0, and 10 pmol of cardiolipin 56:0 was added to each sample. For the analysis of FFAs, 12.5 pmol of arachidonic acid was added to each sample as an internal standard. The samples (1 or 3 nmol) were injected by an autosampler and separated using an ACQUITY UPLC HSS T3 column (Waters, Japan) or SeQuant ZIC-HILIC column (Merck Millipore, Germany). The acquired data from Analyst software (AB Sciex, Japan) were processed with MultiQuant software (AB Sciex, Japan). Lipid peaks were identified according to retention times and multiple reaction monitoring transitions, and quantified relative to the internal standard using the peak area ratio method.

For the measurements of CL and MLCL in Patient 10-derived fibroblasts, fibroblasts were resuspended with 35 µl ice-cold PBS and 5µl of this was transferred to ice-cold RIPA buffer which contained protease inhibitor (1:6.5). The sample in RIPA buffer was lysed for 30 mins. 15 µl aliquot was used for protein quantitation using the DC protein assay (Lowry). The remaining sample in PBS was made up to 980 µl and 20µl of 2 uM CL internal standard was added. A phospholipid extraction using chloroform/methanol was carried out as reported previously (98). The lower organic phase was combined together from two extractions and dried down under nitrogen at 60°C. Once dried, samples were reconstituted in 100 µl methanol and 15 µl was injected onto a Waters Acquity Ultra Performance Liquid Chromatography (UPLC) system (Waters Corporation, UK). Chromatographic settings were based on those previously described (99). Mass spectrometry analysis was carried out on a Waters Xevo TQ-XS Triple Quadrupole Mass Spectrometer (Waters Corp, UK). This was operated in negative ion mode with a source temperature of 150°C and a desolvation gas temperature of 600°C. Nitrogen was used as the desolvation and cone gas, at a flow rate of 1000 and 150 L/Hr respectively. The capillary voltage was maintained at 3.5 kV and the cone voltage was set to 30 V. Mass spectral data was analyzed using Masslynx V4.2 software (Waters Corporation, UK). Mass spectra of all CL and MLCL species was obtained by scanning the m/z range 1000- 1600, between 3.5-7 minutes (scan time 0.2 seconds). Extracted ion chromatograms were then generated for the individual CL and MLCL species and the peak areas were integrated. Analyte values were obtained by calculating the ratio of the CL and MLCL peak areas to the internal standard (C14:0)4 CL peak areas. The values were then standardized to protein levels (Lowry). MLCL made in-house (100), (18:2)4-CL and (C14:0)4 CL standards were used to confirm the retention times of these CL species. Multiple reaction monitoring, using specific parent and daughter ion transitions, was also used to confirm the identity of these species.

### Data availability

Sequencing data have been deposited in the National Center for Biotechnology Information Gene Expression Omnibus (GSE229956).

### Statistics

Quantitative data were generated in at least three independent experiments. All data are expressed as mean ± SD or SEM, as indicated in the figure legends. Significant differences between two groups were assessed by the unpaired Student’s *t* test with Welch’s correction. Other statistical analyses were performed using one-way ANOVA followed by Dunnett’s multiple comparisons. All statistical analyses were done using Graph-pad Prism9 software (GraphPad Software, USA). *P*-values < 0.05 indicated a significant difference between groups. In organoid experiments for quantifying specific area or cell number, *n* represents the number of individual cortical units (the number of organoids tested is specified in figure legends). Organoid and NPC experiments were performed on biological replicates. For each independent experiment, the samples were newly generated from distinct passages of each iPSC line. The measurement of mitochondrial OXPHOS using skin fibroblasts was performed on technical replicates from the Patient 1 sample (*n* = 1).

### Study approval

Genetic and functional analysis using samples from study participants was approved by the institutional review boards at Nagoya City University (Approval Number: 70- 00-0200), Yokohama City University (Approval Number: A170525011), and University College London (Approval Number: #310045/1571740/37/598). Parents and legal guardians of all affected individuals consented to the publication of clinical and genetic information, including video and photographs, and the study was approved by the respective local ethics committees. The generation and application of the iPSCs were approved by the Nagoya University Ethics Committee (Approval Number: 2012-0184) and Nagoya City University Ethics Committee (Approval Number: 19-143). Written informed consent was obtained from patients’ guardians. The analysis using skin fibroblasts derived from the patient with Leigh syndrome was approved by the institutional review board at Jichi Medical University (Approval number: J21-014).

## Author contributions

YN, ISS, YK, and SS designed the study. YN, RM, EK, SM, FM, YO, AS, MM, SA, FA, PH, HSD, CV, MB, AV, AD, MJ, AC, MSZ, HD, SB, MK, CJC, EGK, RAK, TAA, MO, PB, GZ, TS, MZ, CAA, HH, KH, NM, and TT provided clinical care to the study participants, gathered their samples, and performed genetic analyses. YN, MF, MF, ES, and RDSP performed biochemical analyses using patient-derived cells. IO and TM maintained human iPSCs (Windy). YA and NO generated iPSCs derived from Patient 1 and his father. SA, AM, and HO conducted the mitochondrial analysis. MH, YO, and SO performed transcriptomic analysis. TH, YT, and MM performed lipidomic analysis. HT prepared samples and acquired electron microscopy images. YN conducted analyses and generated figures. YN wrote the manuscript with input from ISS, YK, and SS and received feedback and final approval from all authors. ISS, YK, and SS contributed equally to this work.

## Data Availability

All data produced in the present study are available upon reasonable request to the authors.

## Acknowledgments

Human iPSCs (Windy) were kindly provided by Dr Umezawa of the National Center for Child Health and Development (Tokyo, Japan). We thank Dr Padraig J. Flannery and Dr Nina Patel (Neurometabolic Unit, The National Hospital for Neurology and Neurosurgery, London, UK) for their technical assistance in the mass spectrometry analysis. This study was supported by grants from the Japan Society for the Promotion of Science (Grant Numbers JP21463512, JP20K07907, JP21K07869, JP20K21584, and JP21K07803) and the Japan Agency for Medical Research and Development (AMED) under grant numbers, JP21wm0425007, JP20ek0109488, JP22ek0109486, JP22ek0109549, and JP22ek0109493. This study was also supported by grants from the Kawano Masanori Memorial Public Interest Incorporated Foundation for Promotion of Pediatrics, and the Ono Medical Research Foundation to YN, the Daiko Foundation, The Hori Science and Arts Foundation, the Mochida Foundation for Medical and Pharmaceutical Research, the Takeda Science Foundation, iPS Academia Japan Inc., Leave a Nest Co. Ltd. (Ikeda Rika award), and a Grand-in-Aid for research in Nagoya City University (2013009) to ISS, and the Takeda Science Foundation to YK. HSD is supported by the Cologne Clinician Scientist Program/Faculty of Medicine/University of Cologne and funded by the Deutsche Forschungsgemeinschaft (DFG, German Research Foundation, Project No. 413543196) as well as the Koeln Fortune Program / Faculty of Medicine, University of Cologne, (Project No. 371/2021, 243/2022). R.D.S.P. and M.F. are supported by a Medical Research Council (UK) Clinician Scientist Fellowship (MR/S002065/1) and Medical Research Council (UK) award MC_PC_21046 to establish a National Mouse Genetics Network Mitochondria Cluster (MitoCluster). R.D.S.P. is supported by Medical Research Council (UK) strategic award MR/S005021/1 to establish an International Centre for Genomic Medicine in Neuromuscular Diseases (ICGNMD) and is grateful to The Lily Foundation for funding.

## References

1. Florio M, Huttner WB. Neural progenitors, neurogenesis and the evolution of the neocortex. Development. 2014;141(11):2182–2194.

2. Hansen DV, et al. Neurogenic radial glia in the outer subventricular zone of human neocortex. Nature. 2010;464(7288):554–561.

3. Stepien BK, et al. Length of the neurogenic period-a key determinant for the generation of upper-layer neurons during neocortex development and evolution. Front Cell Dev Biol. 2021;9:676911.

4. Wang X, et al. A new subtype of progenitor cell in the mouse embryonic neocortex.Nat Neurosci. 2011;14(5):555–561.

5. Ostrem B, et al. oRGs and mitotic somal translocation - a role in development and disease. Curr Opin Neurobiol. 2017;42:61–67.

6. Mancuso DJ, et al. The genomic organization, complete mRNA sequence, cloning, and expression of a novel human intracellular membrane-associated calcium-independent phospholipase A(2). J Biol Chem. 2000;275(14):9937–9945.

7. Tanaka H, et al. Catalytic residues of group VIB calcium-independent phospholipase A2 (iPLA2gamma). Biochem Biophys Res Commun. 2004;320(4):1284–1290.

8. Tanaka H, et al. A novel Intracellular membrane-bound calcium-independent phospholipase A2. Biochem Biophys Res Commun. 2000;272(2):320–326.

9. Murakami M, et al. Group VIB Ca2+-independent phospholipase A2gamma promotes cellular membrane hydrolysis and prostaglandin production in a manner distinct from other intracellular phospholipases A2. J Biol Chem. 2005;280(14):14028–14041.

10. Mancuso DJ, et al. Complex transcriptional and translational regulation of iPLAgamma resulting in multiple gene products containing dual competing sites for mitochondrial or peroxisomal localization. Eur J Biochem. 2004;271(23-24):4709–4724.

11. Hara S, et al. Calcium-independent phospholipase A2gamma (iPLA2gamma) and its roles in cellular functions and diseases. Biochim Biophys Acta Mol Cell Biol Lipids. 2019;1864(6):861–868.

12. Mancuso DJ, et al. Genetic ablation of calcium-independent phospholipase A2{gamma} leads to alterations in hippocampal cardiolipin content and molecular species distribution, mitochondrial degeneration, autophagy, and cognitive dysfunction. J Biol Chem. 2009;284(51):35632–35644.

13. Mancuso DJ, et al. Genetic ablation of calcium-independent phospholipase A2gamma leads to alterations in mitochondrial lipid metabolism and function resulting in a deficient mitochondrial bioenergetic phenotype. J Biol Chem. 2007;282(48):34611–34622.

14. Yoda E, et al. Mitochondrial dysfunction and reduced prostaglandin synthesis in skeletal muscle of Group VIB Ca2+-independent phospholipase A2gamma-deficient mice. J Lipid Res. 2010;51(10):3003–3015.

15. Chao H, et al. Lowered iPLA2gamma activity causes increased mitochondrial lipid peroxidation and mitochondrial dysfunction in a rotenone-induced model of Parkinson’s disease. Exp Neurol. 2018;300:74–86.

16. Eaddy AC, et al. The role of endoplasmic reticulum Ca2+-independent phospholipase a2gamma in oxidant-induced lipid peroxidation, Ca2+ release, and renal cell death. Toxicol Sci. 2012;128(2):544–552.

17. Liu GY, et al. The phospholipase iPLA2gamma is a major mediator releasing oxidized aliphatic chains from cardiolipin, integrating mitochondrial bioenergetics and signaling. J Biol Chem. 2017;292(25):10672–10684.

18. Sharma J, et al. The absence of myocardial calcium-independent phospholipase A2gamma results in impaired prostaglandin E2 production and decreased survival in mice with acute Trypanosoma cruzi infection. Infect Immun. 2013;81(7):2278–2287.

19. Sharma J, McHowat J. PGE2 release from tryptase-stimulated rabbit ventricular myocytes is mediated by calcium-independent phospholipase A2gamma. Lipids. 2011;46(5):391–397.

20. Harmouch F, et al. PNPLA 8 mutation in mitochondrial disease: Second case worldwide. Acta Scientific Clinical Case Reports. 2020;1(10):19–21.

21. Masih S, et al. Homozygous missense variation in PNPLA8 causes prenatal-onset severe neurodegeneration. Mol Syndromol. 2021;12(3):174–178.

22. Saunders CJ, et al. Loss of function variants in human PNPLA8 encoding calcium-independent phospholipase A2 gamma recapitulate the mitochondriopathy of the homologous null mouse. Hum Mutat. 2015;36(3):301–306.

23. Shukla A, et al. A neurodegenerative mitochondrial disease phenotype due to biallelic loss-of-function variants in PNPLA8 encoding calcium-independent phospholipase A2gamma. Am J Med Genet A. 2018;176(5):1232–1237.

24. Sherry ST, et al. dbSNP: the NCBI database of genetic variation. Nucleic Acids Res. 2001;29(1):308–311.

25. Fairley S, et al. The International Genome Sample Resource (IGSR) collection of open human genomic variation resources. Nucleic Acids Res. 2020;48(D1):D941–D947.

26. Tadaka S, et al. jMorp updates in 2020: large enhancement of multi-omics data resources on the general Japanese population. Nucleic Acids Res. 2021;49(D1):D536–D544.

27. Karczewski KJ, et al. The mutational constraint spectrum quantified from variation in 141,456 humans. Nature. 2020;581(7809):434–443.

28. Jaganathan K, et al. Predicting splicing from primary sequence with deep learning. Cell. 2019;176(3):535–548 e524.

29. Chicco AJ, Sparagna GC. Role of cardiolipin alterations in mitochondrial dysfunction and disease. Am J Physiol Cell Physiol. 2007;292(1):C33–44.

30. Nishino K, et al. Defining hypo-methylated regions of stem cell-specific promoters in human iPS cells derived from extra-embryonic amnions and lung fibroblasts. PLoS One. 2010;5(9):e13017.

31. Iwao T, et al. Differentiation of human induced pluripotent stem cells into functional enterocyte-like cells using a simple method. Drug Metab Pharmacokinet. 2014;29(1):44–51.

32. Kondo Y, et al. Histone deacetylase inhibitor valproic acid promotes the differentiation of human induced pluripotent stem cells into hepatocyte-like cells. PLoS One. 2014;9(8):e104010.

33. Lancaster MA, et al. Cerebral organoids model human brain development and microcephaly. Nature. 2013;501(7467):373–379.

34. Pasca AM, et al. Functional cortical neurons and astrocytes from human pluripotent stem cells in 3D culture. Nat Methods. 2015;12(7):671–678.

35. Sloan SA, et al. Generation and assembly of human brain region-specific three-dimensional cultures. Nat Protoc. 2018;13(9):2062–2085.

36. Xiang Y, et al. hESC-derived thalamic organoids form reciprocal projections when fused with cortical organoids. Cell Stem Cell. 2019;24(3):487–497 e487.

37. Di Lullo E, Kriegstein AR. The use of brain organoids to investigate neural development and disease. Nat Rev Neurosci. 2017;18(10):573–584.

38. Martinez-Cerdeno V, Noctor SC. Neural Progenitor Cell Terminology. Front Neuroanat. 2018;12:104.

39. Kalebic N, et al. Neocortical expansion due to increased proliferation of basal progenitors is linked to changes in their morphology. Cell Stem Cell. 2019;24(4):535–550 e539.

40. Kostic M, et al. YAP activity Is necessary and sufficient for basal progenitor abundance and proliferation in the developing neocortex. Cell Rep. 2019;27(4):1103–1118 e1106.

41. Homem CC, et al. Proliferation control in neural stem and progenitor cells. Nat Rev Neurosci. 2015;16(11):647–659.

42. Severino M, et al. Definitions and classification of malformations of cortical development: practical guidelines. Brain. 2020;143(10):2874–2894.

43. Uzquiano A, et al. Cortical progenitor biology: key features mediating proliferation versus differentiation. J Neurochem. 2018;146(5):500–525.

44. Zhang W, et al. Cerebral organoid and mouse models reveal a RAB39b-PI3K-mTOR pathway-dependent dysregulation of cortical development leading to macrocephaly/autism phenotypes. Genes Dev. 2020;34(7-8):580–597.

45. Ohnuki M, et al. Generation and characterization of human induced pluripotent stem cells. Curr Protoc Stem Cell Biol. 2009;Chapter 4:Unit 4A 2.

46. Zhao J, et al. APOE4 exacerbates synapse loss and neurodegeneration in Alzheimer’s disease patient iPSC-derived cerebral organoids. Nat Commun. 2020;11(1):5540.

47. Bershteyn M, et al. Human iPSC-derived cerebral organoids model cellular features of lissencephaly and reveal prolonged mitosis of outer radial glia. Cell Stem Cell. 2017;20(4):435–449 e434.

48. Khacho M, et al. Mitochondria as central regulators of neural stem cell fate and cognitive function. Nat Rev Neurosci. 2019;20(1):34–48.

49. Malatesta P, et al. Radial glia and neural stem cells. Cell Tissue Res. 2008;331(1):165–178.

50. Tomooka Y, et al. Reconstruction of neural tube-like structures in vitro from primary neural precursor cells. Proc Natl Acad Sci U S A. 1993;90(20):9683–9687.

51. Honda M, et al. Photo-isolation chemistry for high-resolution and deep spatial transcriptome with mouse tissue sections. STAR Protoc. 2022;3(2):101346.

52. Honda M, et al. High-depth spatial transcriptome analysis by photo-isolation chemistry. Nat Commun. 2021;12(1):4416.

53. Maere S, et al. BiNGO: a Cytoscape plugin to assess overrepresentation of gene ontology categories in biological networks. Bioinformatics. 2005;21(16):3448–3449.

54. Liberzon A, et al. The Molecular Signatures Database (MSigDB) hallmark gene set collection. Cell Syst. 2015;1(6):417–425.

55. Liberzon A, et al. Molecular signatures database (MSigDB) 3.0. Bioinformatics. 2011;27(12):1739–1740.

56. Subramanian A, et al. Gene set enrichment analysis: a knowledge-based approach for interpreting genome-wide expression profiles. Proc Natl Acad Sci U S A. 2005;102(43):15545–15550.

57. Florio M, et al. Human-specific gene ARHGAP11B promotes basal progenitor amplification and neocortex expansion. Science. 2015;347(6229):1465–1470.

58. Merico D, et al. Enrichment map: a network-based method for gene-set enrichment visualization and interpretation. PLoS One. 2010;5(11):e13984.

59. Bretones G, et al. Myc and cell cycle control. Biochim Biophys Acta. 2015;1849(5):506–516.

60. Zinin N, et al. MYC proteins promote neuronal differentiation by controlling the mode of progenitor cell division. EMBO Rep. 2014;15(4):383–391.

61. Fan X, et al. Single-cell transcriptome analysis reveals cell lineage specification in temporal-spatial patterns in human cortical development. Sci Adv. 2020;6(34):eaaz2978.

62. Iwata R, Vanderhaeghen P. Regulatory roles of mitochondria and metabolism in neurogenesis. Curr Opin Neurobiol. 2021;69:231–240.

63. Khacho M, Slack RS. Mitochondrial dynamics in the regulation of neurogenesis: From development to the adult brain. Dev Dyn. 2018;247(1):47–53.

64. Schaefer CF, et al. PID: the Pathway Interaction Database. Nucleic Acids Res. 2009;37(Database issue):D674-679.

65. Geraldo LHM, et al. Role of lysophosphatidic acid and its receptors in health and disease: novel therapeutic strategies. Signal Transduct Target Ther. 2021;6(1):45.

66. Noguchi K, et al. Lysophosphatidic acid (LPA) and its receptors. Curr Opin Pharmacol. 2009;9(1):15–23.

67. Kingsbury MA, et al. Non-proliferative effects of lysophosphatidic acid enhance cortical growth and folding. Nat Neurosci. 2003;6(12):1292–1299.

68. Supek F, et al. To NMD or not to NMD: Nonsense-mediated mRNA decay in cancer and other genetic diseases. Trends Genet. 2021;37(7):657–668.

69. Baker KE, Parker R. Nonsense-mediated mRNA decay: terminating erroneous gene expression. Curr Opin Cell Biol. 2004;16(3):293–299.

70. Nowakowski TJ, et al. Transformation of the radial glia scaffold demarcates two stages of human cerebral cortex development. Neuron. 2016;91(6):1219–1227.

71. Yu TW, et al. Mutations in WDR62, encoding a centrosome-associated protein, cause microcephaly with simplified gyri and abnormal cortical architecture. Nat Genet. 2010;42(11):1015–1020.

72. Ghosh L, Jessberger S. Supersize me-new insights into cortical expansion and gyration of the mammalian brain. EMBO J. 2013;32(13):1793–1795.

73. Ostrem BE, et al. Control of outer radial glial stem cell mitosis in the human brain. Cell Rep. 2014;8(3):656–664.

74. Pollen AA, et al. Molecular identity of human outer radial glia during cortical development. Cell. 2015;163(1):55–67.

75. Saade M, et al. A centrosomal view of CNS growth. Development. 2018;145(21):dev170613.

76. Murakami M, et al. Updating phospholipase A(2) biology. Biomolecules. 2020;10(10):1457.

77. Shindou H, Shimizu T. Acyl-CoA:lysophospholipid acyltransferases. J Biol Chem. 2009;284(1):1–5.

78. Pinson A, et al. Human TKTL1 implies greater neurogenesis in frontal neocortex of modern humans than Neanderthals. Science. 2022;377(6611):eabl6422.

79. Denomme-Pichon AS, et al. Accelerated genome sequencing with controlled costs for infants in intensive care units: a feasibility study in a French hospital network. Eur J Hum Genet. 2022;30(5):567–576.

80. Chou JH, et al. PediTools electronic growth chart calculators: Applications in clinical care, research, and quality improvement. J Med Internet Res. 2020;22(1):e16204.

81. Kuczmarski R, et al. CDC Growth Charts for the United States: methods and development. Vital Health Stat 11. 2002;246:1–190.

82. Efthymiou S, et al. Biallelic mutations in neurofascin cause neurodevelopmental impairment and peripheral demyelination. Brain. 2019;142(10):2948–2964.

83. Kato K, et al. MYCN de novo gain-of-function mutation in a patient with a novel megalencephaly syndrome. J Med Genet. 2019;56(6):388–395.

84. Saida K, et al. Brain monoamine vesicular transport disease caused by homozygous SLC18A2 variants: A study in 42 affected individuals. Genet Med. 2023;25(1):90–102.

85. Schuster S, et al. Generation of a homozygous CRISPR/Cas9-mediated knockout human iPSC line for the STUB1 locus. Stem Cell Res. 2019;34:101378.

86. Miyazaki T, et al. Efficient adhesion culture of human pluripotent stem cells using laminin fragments in an uncoated manner. Sci Rep. 2017;7:41165.

87. Arioka Y, et al. Induced pluripotent stem cells derived from a schizophrenia patient with ASTN2 deletion. Stem Cell Res. 2018;30:81–84.

88. Arioka Y, et al. Single-cell trajectory analysis of human homogenous neurons carrying a rare RELN variant. Transl Psychiatry. 2018;8(1):129.

89. Mack AA, et al. Generation of induced pluripotent stem cells from CD34+ cells across blood drawn from multiple donors with non-integrating episomal vectors. PLoS One. 2011;6(11):e27956.

90. Okita K, et al. A more efficient method to generate integration-free human iPS cells. Nat Methods. 2011;8(5):409–412.

91. Arioka Y, et al. Behavior of leucine-rich repeat-containing G-protein coupled receptor 5-expressing cells in the reprogramming process. Stem Cell Res. 2017;20:1–9.

92. Sakaguchi H, et al. Generation of functional hippocampal neurons from self-organizing human embryonic stem cell-derived dorsomedial telencephalic tissue. Nat Commun. 2015;6:8896.

93. Shimada IS, et al. Self-renewal and differentiation of reactive astrocyte-derived neural stem/progenitor cells isolated from the cortical peri-infarct area after stroke. J Neurosci. 2012;32(23):7926–7940.

94. Meharena HS, et al. Down-syndrome-induced senescence disrupts the nuclear architecture of neural progenitors. Cell Stem Cell. 2022;29(1):116–130 e117.

95. Tanaka Y, et al. LPIAT1/MBOAT7 depletion increases triglyceride synthesis fueled by high phosphatidylinositol turnover. Gut. 2021;70(1):180–193.

96. Bligh EG, Dyer WJ. A rapid method of total lipid extraction and purification. Can J Biochem Physiol. 1959;37(8):911–917.

97. Bartlett GR. Phosphorus assay in column chromatography. Journal of Biological Chemistry. 1959;234(3):466–468.

98. Folch J, et al. A simple method for the isolation and purification of total lipides from animal tissues. Journal of Biological Chemistry. 1957;226(1):497–509.

99. Vaz FM, et al. An improved functional assay in blood spot to diagnose Barth syndrome using the monolysocardiolipin/cardiolipin ratio. J Inherit Metab Dis. 2022;45(1):29–37.

100. Kim J, Hoppel CL. Monolysocardiolipin: improved preparation with high yield. J Lipid Res. 2011;52(2):389–392.

