## Supplemental Material for "Biallelic null variants in *PNPLA8* cause microcephaly through the reduced abundance of basal radial glia"

### Supplemental text

#### Case reports

##### Family 1

This case report has been removed from the preprint version to comply with medrxiv policy. Please see the published version or contact the authors if you are interested in this information.

##### Family 2

This case report has been removed from the preprint version to comply with medrxiv policy. Please see the published version or contact the authors if you are interested in this information.

##### Family 3

This case report has been removed from the preprint version to comply with medrxiv policy. Please see the published version or contact the authors if you are interested in this information.

##### Family 4

This case report has been removed from the preprint version to comply with medrxiv policy. Please see the published version or contact the authors if you are interested in this information.

##### Family 5

This case report has been removed from the preprint version to comply with medrxiv policy. Please see the published version or contact the authors if you are interested in this information.

##### Family 6

This case report has been removed from the preprint version to comply with medrxiv policy. Please see the published version or contact the authors if you are interested in this information.

##### Family 7

This case report has been removed from the preprint version to comply with medrxiv policy. Please see the published version or contact the authors if you are interested in this information.

##### Family 8

This case report has been removed from the preprint version to comply with medrxiv policy. Please see the published version or contact the authors if you are interested in this information.

##### Family 9

This case report has been removed from the preprint version to comply with medrxiv policy. Please see the published version or contact the authors if you are interested in this information.

##### Family 10

This case report has been removed from the preprint version to comply with medrxiv policy. Please see the published version or contact the authors if you are interested in this information.

#### Supplemental Methods

*Cell lines.* EBV-induced lymphoblastoid cell lines (LCLs) were established from peripheral blood using a standard method (3). Briefly, peripheral blood was collected in a tube. Ficoll was added 1:1, followed by centrifugation (1,500 rpm, 30 min). The layer containing mononuclear cells was separated into another tube, followed by centrifugation (1,500 rpm, 5 min) and discarding the supernatant. We resuspended the cells in 2.5 mL of RPMI-1640 supplemented with 20% FBS and 2.4 μg/mL cyclosporin A, and subsequently infected with EBV. Cells were cultured in RPMI-1640 supplemented with 10% FBS, 100 U/mL penicillin, 100 mg/mL streptomycin, and 29.2 mg/mL L-glutamine for 30 more days before collection of LCLs. Skin fibroblasts derived from Patient 1 were established from skin punch biopsies using a standard method (4). In brief, tissue pieces were explanted to cell culture plates and cultured in DMEM with 10% FBS, 100 U/mL penicillin, 100 mg/mL streptomycin, and 29.2 mg/mL L-glutamine until fibroblast outgrowth. The fibroblasts were passaged for expansion but maintained at a low passage under mycoplasma-free conditions. For control experiments, normal human skin fibroblasts were purchased (Control 1: lot# 428Z014.2, Control 2: lot# 425Z026.3) from PromoCell or obtained from MRC Centre for Neuromuscular Disorders Biobank London. All cell lines were maintained at 37°C and 5% CO2.

*TA cloning.* Total RNA was extracted using an RNeasy mini kit from Patient 1 LCLs for reverse transcription PCR (RT-PCR). cDNAs were generated from RNAs templates using SuperScript IV reverse transcriptase. PCR was then performed using Taq DNA polymerase to leave a 3' A-overhang. The obtained RT-PCR fragments were subcloned into PCR-TOPO vector using a TOPO TA Cloning kit and subsequently transformed into chemically competent *E. coli* cells according to the manufacturer's instructions. Individual clones were grown and picked via blue/white color selection of colonies mediated by β-galactosidase expression. Plasmids containing inserts were purified using a Miniprep kit and sequenced by Genetic Analyzer (SeqStudio; Applied Biosystems, USA). Primer sequences are provided in Supplemental Table 3.

*RNA isolation and gene expression analysis.* For the gene expression analysis of Patient 10-derived cells, total RNA was isolated from fibroblasts, using the RNeasy Mini Kit, and genomic DNA contamination was removed using DNA-free DNA Removal kit. Quality of the extracted RNA was assessed by 1% agarose gel electrophoresis and from the A260nm/A280nm absorbance ratio (Nanodrop One; ThermoFisher Scientific, USA) and cDNA was synthesized using the High-Capacity cDNA Kit, according to the manufacturer's instructions. The expression of *PNPLA8* mRNA was measured by quantitative PCR (qPCR). Gene expression was determined using TaqMan Fast Advance Master Mix according to the manufacturer's protocol and qPCR reactions were carried out on a QuantStudio 5 thermal cycler (ThermoFisher Scientific, USA). The human probes PNPLA8 (Hs.PT.58.39503048) and the housekeeping β2-microglobulin were from Integrated DNA Technologies.

*Immunoblotting.* To obtain protein extracts for immunoblotting, cells in a 6-well dish were rinsed twice with ice-cold PBS and lysed immediately with RIPA lysis buffer containing 1% protease inhibitor cocktail. The lysate was centrifuged (1,5000 × g, 4°C) for 30 min, and the supernatant was eluted with sodium dodecyl sulfate (SDS) buffer (0.125 mol/L Tris–HCl, pH 6.8, 10% glycerol, 4% SDS, 0.01% bromophenol blue, 5% 2-mercaptoethanol). After boiling at 100°C for 5 min, whole-cell lysates were subjected to SDS-polyacrylamide gel electrophoresis, followed by transfer to polyvinylidene difluoride membranes using a powerpack HC (Bio-Rad, USA). Non-specific binding was blocked with 5% skim milk in 0.1% PBS-Tween20 for 1 h at RT, and proteins were probed with primary antibodies diluted in 0.1% PBS-Tween20 at 4°C overnight. After washing with 0.1% PBS-Tween20, the horseradish peroxidase-labeled secondary antibodies against rabbit or mouse IgG were probed for 1 h at RT. The ECL prime HRP detection kit was used to detect the signal. All images were acquired from an Amersham Imager 600 (GE Healthcare, USA). Subsequently, the blot was treated with Western Blot Stripping Buffer for 15 min. The same blot was reprobed with an anti-β-actin antibody as an internal control. Primary and secondary antibodies are listed in Supplemental Table 3.

For the analysis of PNPLA8 protein expression in Patient 10-derived fibroblasts, total protein extracts were resolved on a 10-12% Tricine polyacrylamide gel, transferred onto Trans-Blot nitrocellulose membrane, then incubated with primary antibodies against the following proteins: PNPLA8 and GAPDH followed by Infrared dye-labeled secondary antibodies. The antibodies used are listed in Supplemental Table 3. Images were detected with the Odyssey CLx infrared imager (Li-Cor, USA) at 680 and 800 nm.

*Transmission electron microscopy.* Cells were fixed in 2% glutaraldehyde in 0.1 M phosphate buffer (pH 7.4) at 4°C overnight, and post-fixed in 2% osmium tetroxide in 0.1 M phosphate buffer (pH 7.4) at 4°C for 45 min. The cells were then dehydrated with a graded ethanol series and embedded in Quetol 812 epoxy resin at 60°C for 48 h. Ultrathin sections 80-100 nm thick were obtained with an Ultracut-UCT (Leica, Germany), stained with uranyl acetate for 15 min, and stained with modified Sato's lead solution for 5 min (5). Transmission electron microscopy observations were performed using an electron microscope (JEM-1400 Plus; JEOL, Japan).

*Measuring mitochondrial function.* The oxygen consumption ratio (OCR) of cells was measured using an extracellular flux analyzer (Seahorse XFe 96; Agilent Technologies, USA) as previously described (6). We seeded 20,000 cells into XFe96-cell culture plates and cultured them in media for skin fibroblasts and iPSC-derived NPCs, respectively. Before the analysis, the culture medium was replaced with Seahorse XF DMEM Medium containing 10 mM glucose, 1 mM pyruvate, and 2 mM glutamine. Subsequently, cells were incubated without $CO_2$ at 37°C for 1 h. We examined 15 wells per experiment. Following the analysis, the DNA content was examined with a CyQUANT Cell Proliferation Assay Kit. We used the normalization function of an Agilent Seahorse XF assay to account for variations in DNA content. Basal OCR, ATP-linked OCR, maximal OCR, and spare capacity were determined by measuring the OCR after the sequential injection of oligomycin (to inhibit ATP synthase), carbonyl cyanide p-trifluoromethoxy-phenyl-hydrazone (FCCP; allowing for maximum electron flux by uncoupling oxidative phosphorylation), and rotenone (to inhibit complex I), respectively. As a positive control, skin fibroblasts derived from a patient with Leigh syndrome (complex I deficiency) were used.

**Supplemental References**


1. Denomme-Pichon AS, et al. Accelerated genome sequencing with controlled costs for infants in intensive care units: a feasibility study in a French hospital network. *Eur J Hum Genet*. 2022;30(5):567-576.
2. Harmouch F, et al. PNPLA 8 mutation in mitochondrial disease: Second case worldwide. *Acta Scientific Clinical Case Reports*. 2020;1(10):19-21.
3. Neitzel H. A routine method for the establishment of permanent growing lymphoblastoid cell lines. *Hum Genet*. 1986;73(4):320-326.
4. Rittie L, Fisher GJ. Isolation and culture of skin fibroblasts. *Methods Mol Med*. 2005;117:83-98.
5. Hanaichi T, et al. A stable lead by modification of Sato's method. *J Electron Microsc (Tokyo)*. 1986;35(3):304-306.
6. Miyauchi A, et al. Apomorphine rescues reactive oxygen species-induced apoptosis of fibroblasts with mitochondrial disease. *Mitochondrion*. 2019;49:111-120.

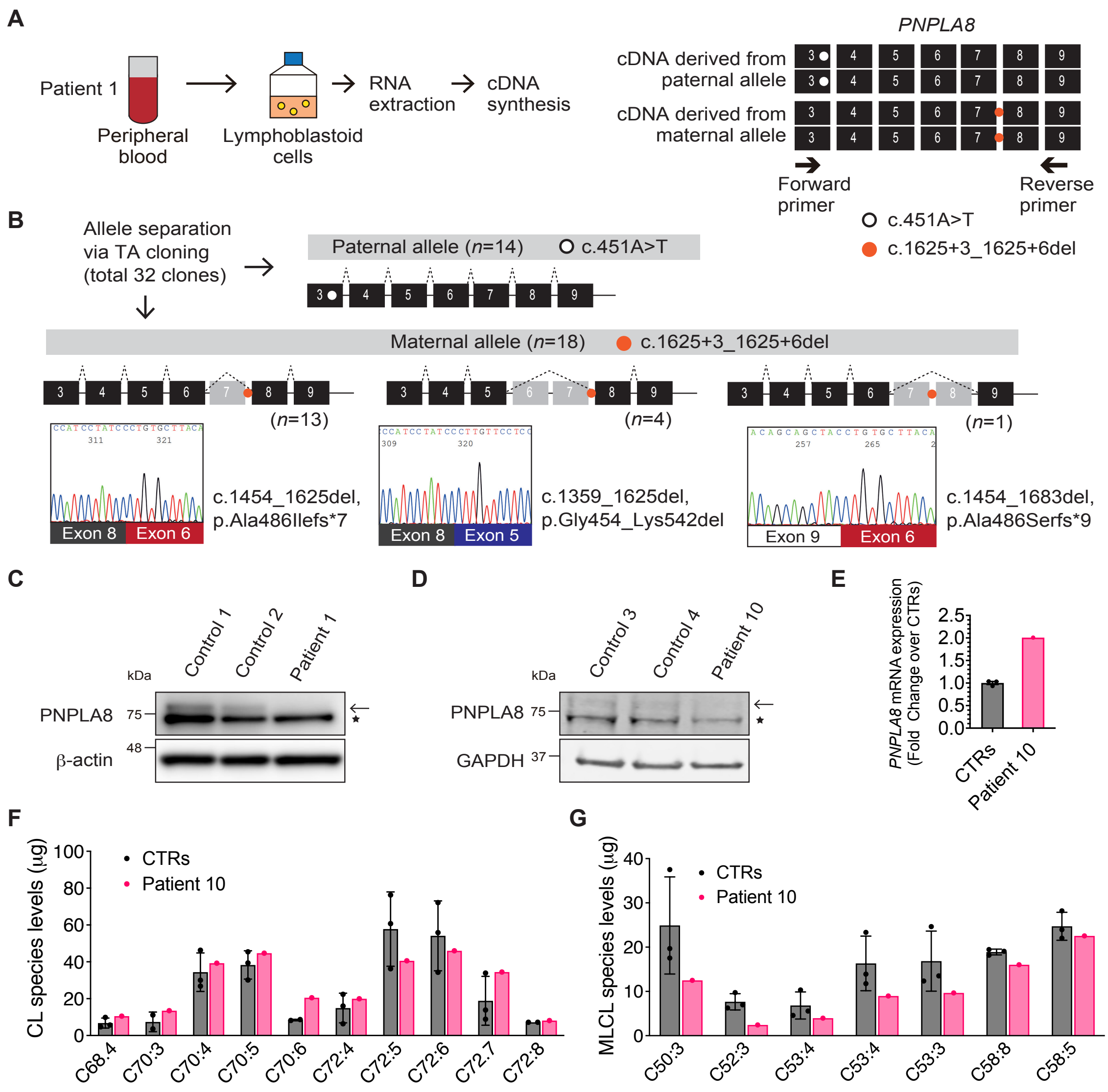


**Supplemental Figure 1. Functional characterization of *PNPLA8* variants using patient-derived cells.** (**A**) Overview of the protocol for RT-PCR. PCR primers were designed to include both paternally- and maternally-derived variants. (**B**) Sanger sequencing of the 32 clones obtained from TA cloning. All transcripts from the maternal allele distinguished by the absence of the nonsense variant in exon 3 showed multiple aberrant splicing patterns, such as skipping of exon 7 ($n = 13$), exon 6-7 ($n = 4$), and exon 7-8 ($n = 1$). Electropherograms represent antisense strands. (**C** and **D**) Immunoblotting analysis of PNPLA8 (MW of 77 kDa) in control and patient-derived skin fibroblasts from Family 1 (**C**) and 10 (**D**). β-actin (42 kDa) or GAPDH (36 kDa) was used as an internal protein loading control. The arrow indicates the 77 kDa PNPLA8 band, and the asterisk indicates a non-specific band. Full-length blots are shown in Supplemental Figures 8A and B. (**E**) Quantitative PCR analysis of *PNPLA8* transcripts in Patient 10-derived skin fibroblasts. Regarding the controls (CTRs), average values ± SD from three samples are plotted. (**F** and **G**) Cardiolipin (CL) and monolysocardiolipin (MLCL) acyl-chain composition profiles of Patient 10-derived skin fibroblasts. Regarding the CTRs, average values ± SD from three samples are plotted.

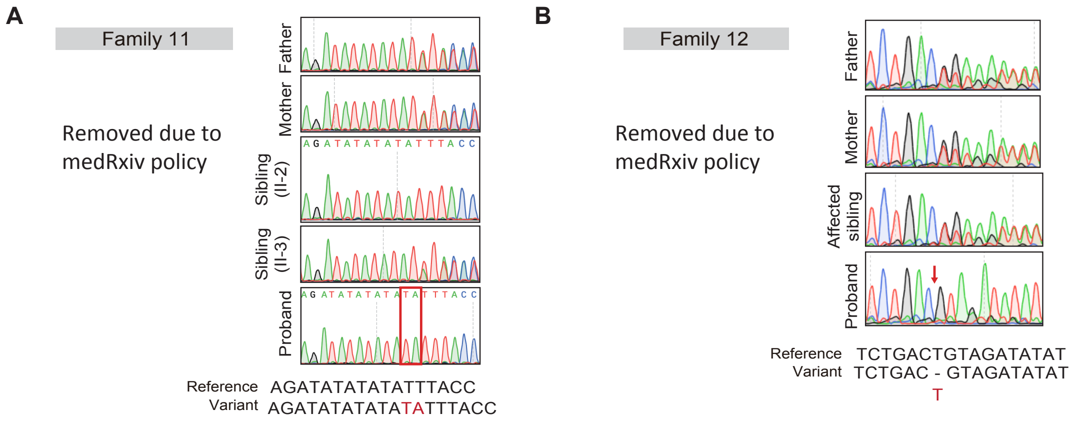


**Supplemental Figure 2. Genetic data for non-neuronal cases.** (**A** and **B**) Family pedigree and Sanger sequencing of the variants. Altered sequences are shown in *red*. The probands are indicated by arrows. Double lines, first cousin status; squares, males; circles, females; black fills, affected individuals. Variants in *PNPLA8* are denoted as 'mut', and WT sequences in *PNPLA8* are represented as 'wt'.

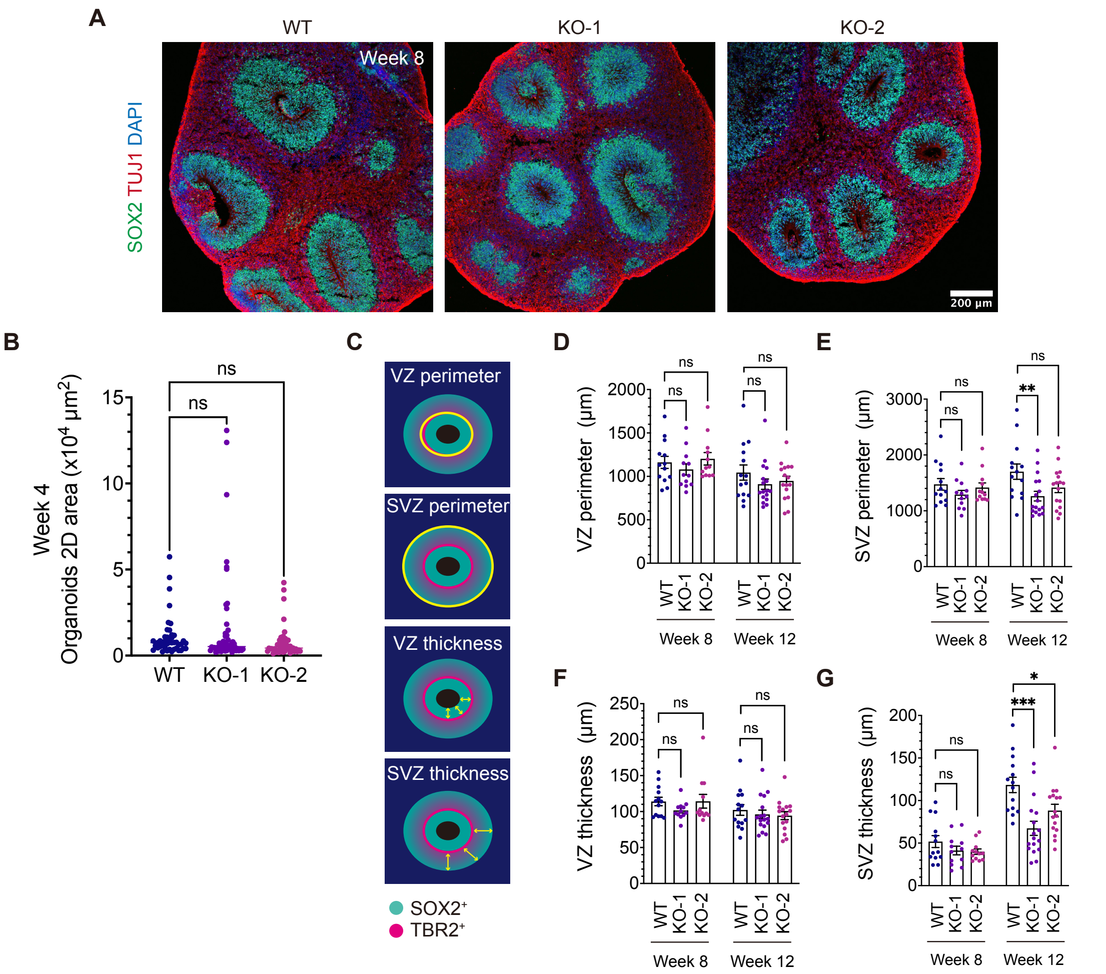


**Supplemental Figure 3. Quantification of the size of cerebral organoids.** (**A**) Representative immunofluorescence images of $SOX2^+$ NPCs and $TUJ1^+$ neurons in cerebral organoids at eight weeks of culture. Scale bar, 200 μm. (**B**) Quantification of the organoid area at four weeks of culture. Average values ± SEM from six independent experiments (at least five organoids per experiment) are plotted. WT ($n$ = 44); KO-1 ($n$ = 53); KO-2 ($n$ = 47). $n$ represents the number of organoids. ns, not significant. One-way ANOVA followed by Dunnett's multiple comparisons test. (**C**) Schematic illustration of regions of the VZ and SVZ according to the spatial distribution of NPCs. The thickness of the VZ and SVZ was determined by the average of three measurements. (**D-G**) Quantification of the perimeter and the thickness at eight and 12 weeks of culture. Average values ± SEM from three independent experiments (at least three organoids per experiment) are plotted. *$P < 0.05$, **$P < 0.01$, ***$P < 0.001$; ns, not significant. One-way ANOVA followed by Dunnett's multiple comparisons test for each time point.

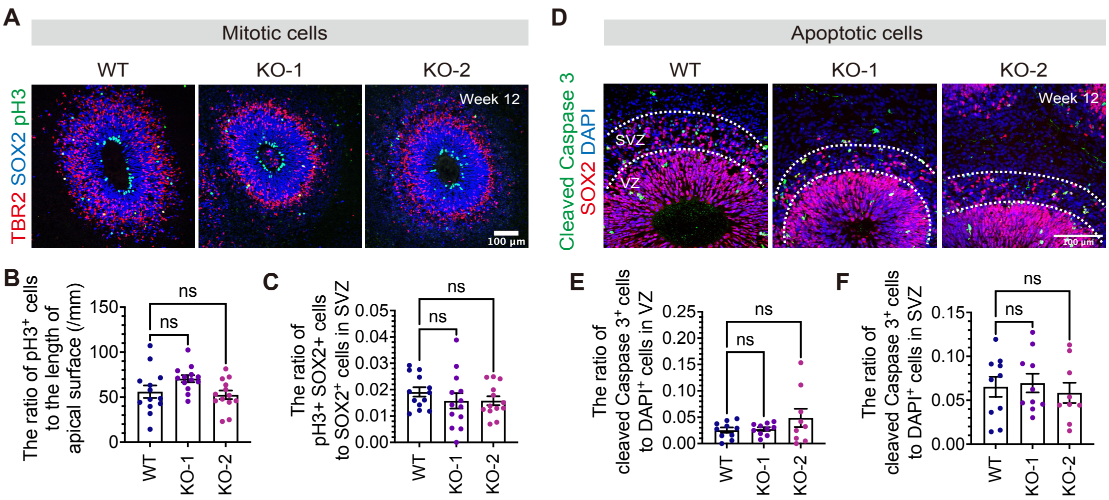


**Supplemental Figure 4. The estimation of mitosis and apoptosis in *PNPLA8* KO cerebral organoids.** (**A**) Representative immunofluorescence images of pH3$^+$ mitotic cells, SOX2$^+$ NPCs, and TBR2$^+$ bIPs at 12 weeks of culture. Scale bar, 100 µm. (**B** and **C**) Quantification of pH3$^+$ cells in the apical surface (**B**) and SVZ (**C**). The apical surface and SVZ are defined according to the spatial distribution of aRGCs and bIPs. Average values ± SEM from three independent experiments (at least three organoids per experiment) are plotted. WT ($n$ = 13); KO-1 ($n$ = 13); KO-2 ($n$ = 13). ns, not significant. One-way ANOVA followed by Dunnett's multiple comparisons test. (**D**) Representative immunofluorescence images of SOX2$^+$ aRGCs, and Cleaved Caspase 3$^+$ apoptotic cells at 12 weeks of culture. Scale bar, 100 µm. (**E** and **F**) Quantification of Cleaved Caspase 3$^+$ cells in the VZ (**E**) and SVZ (**F**). The VZ and SVZ are defined according to the spatial distribution of aRGCs. Average values ± SEM from three independent experiments (at least three organoids per experiment) are plotted. WT ($n$ = 9); KO-1 ($n$ = 9); KO-2 ($n$ = 10). ns, not significant. One-way ANOVA followed by Dunnett's multiple comparisons test.

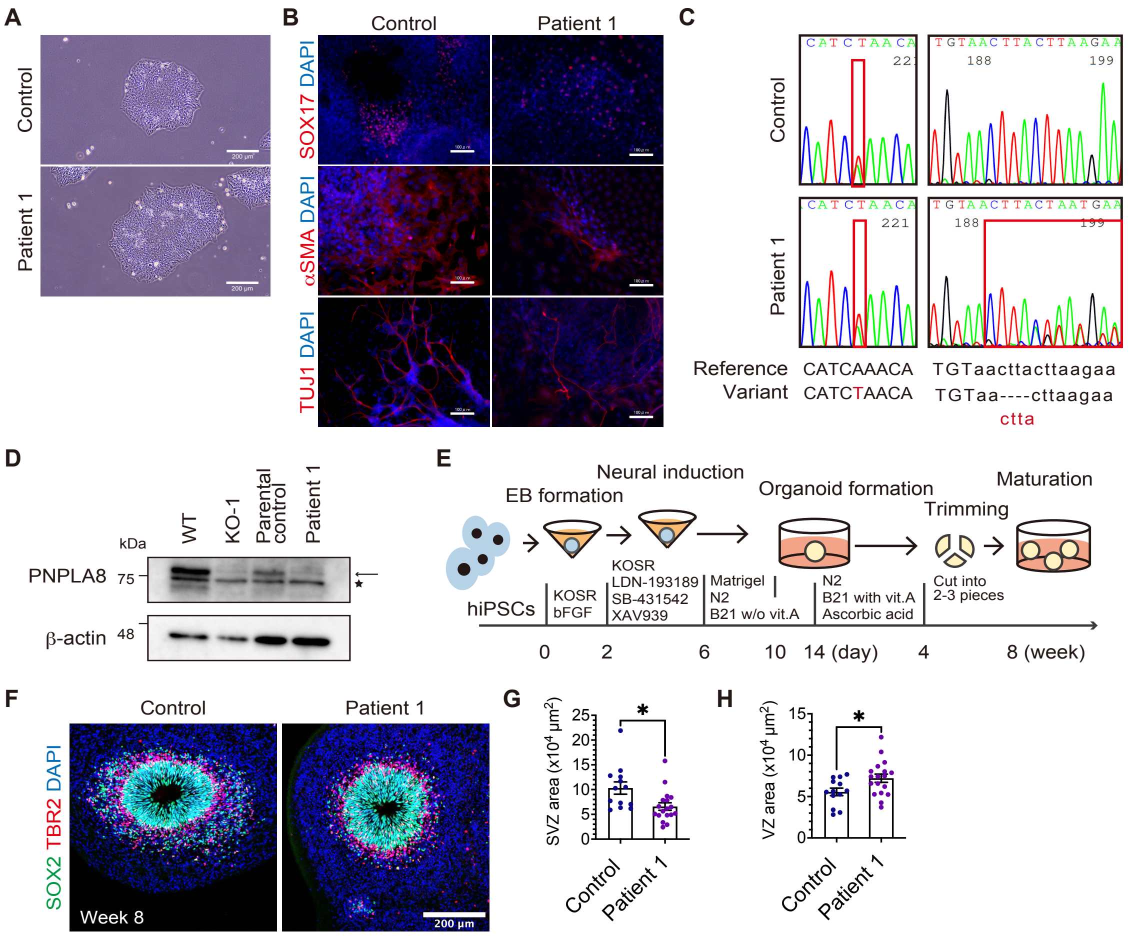


**Supplemental Figure 5. Characterization of iPSCs and cerebral organoids generated from patients with pathogenic *PNPLA8* variants.** (**A**) Representative bright-field images of iPSCs derived from Patient 1 and the parental control. Scale bars, 200 µm. (**B**) Representative immunofluorescence images of $SOX17^+$ endoderm cells, $\alpha SMA^+$ mesoderm cells, and $TUJ1^+$ ectoderm cells. Scale bars, 100 µm. (**C**) Sanger sequencing of the variants in compound heterozygous state in patient iPSCs and heterozygous state in control iPSCs. Altered sequences in exons (upper-case letters) and introns (lower-case letters) are shown in red. (**D**) Immunoblotting analysis of PNPLA8 levels in iPSC lines derived from Patient 1 and parental control with the heterozygous nonsense variant in *PNPLA8*. WT and *PNPLA8* KO-1 iPSCs were used as positive and negative controls, respectively. β-actin was used as an internal protein loading control. The arrow indicates the 77 kDa PNPLA8 band, and the asterisk indicates a non-specific band. Full-length blots are shown in Supplemental Figure 8D. (**E**) Schematic representation of the experimental workflow for the generation of cerebral organoids. (**F**) Representative immunofluorescence images of $SOX2^+$ aRGCs and $TBR2^+$ bIPs at 8 weeks of culture. Scale bars, 200 µm. (**G** and **H**) Quantification of the 2D area of the VZ

(**G**) and SVZ (**H**). Average values ± SEM from the number of organoids in three independent experiments (at least three organoids per experiment) are plotted. control (*n* = 13); Patient 1 (*n* = 18); **P* < 0.05. Unpaired Student's *t* test with Welch's correction.

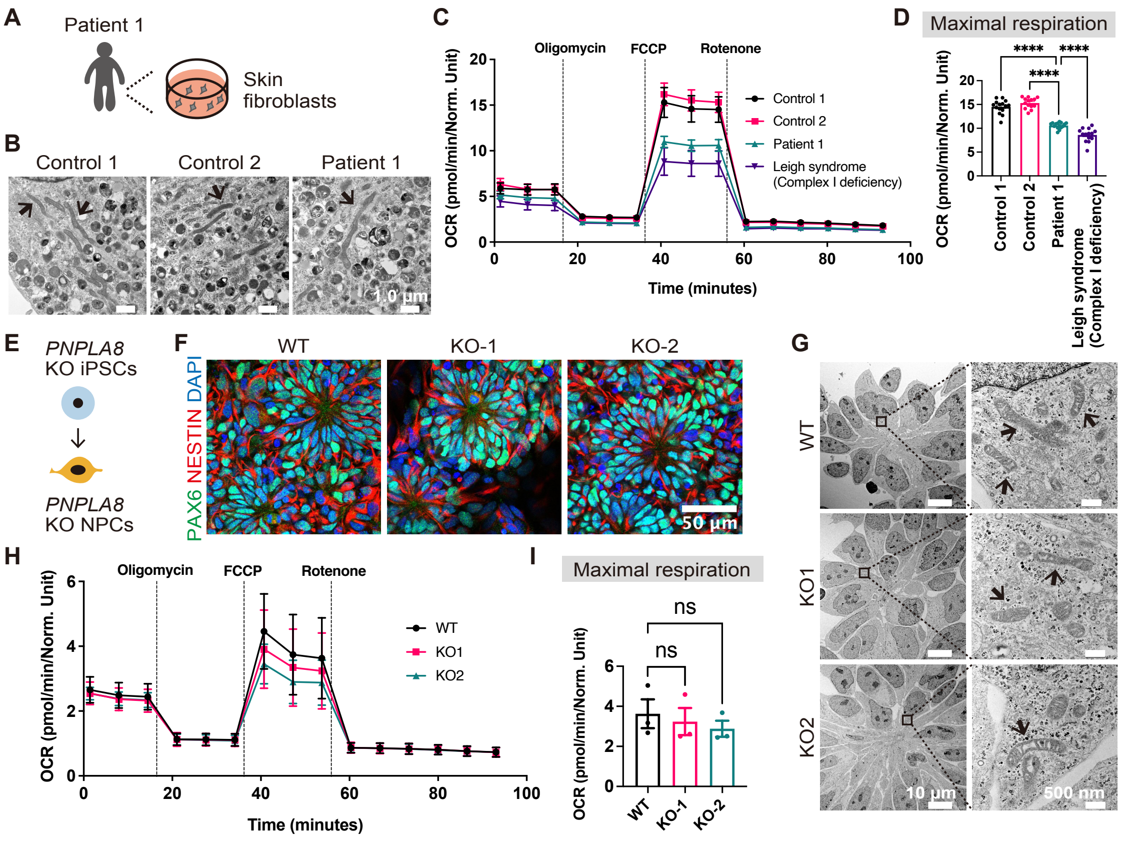


**Supplemental Figure 6. Role of PNPLA8 in the regulation of mitochondrial function and morphology.** (**A**) Skin fibroblasts were generated from Patient 1. (**B**) Transmission electron microscopy (TEM) images of control and patient-derived skin fibroblasts. Black arrows indicate mitochondria. Scale bars, 1 μm. (**C**) Oxygen consumption rate (OCR) measurement by an extracellular flux analyzer in skin fibroblasts. (**D**) Quantification of the maximal OCR after injection of carbonyl cyanide p-trifluoromethoxy-phenyl-hydrazone (FCCP). Average values ± SEM from 14 or 15 technical replicate experiments are plotted. Control 1 ($n$ = 15); control 2 ($n$ = 14); Patient 1 ($n$ = 15); Leigh syndrome ($n$ = 14). ****$P < 0.0001$. One-way ANOVA followed by Dunnett's multiple comparisons test. (**E**) Schematic illustration of the generation of iPSC-derived NPCs. (**F**) Representative immunofluorescence images of $PAX6^{+}$ $NESTIN^{+}$ NPCs generated from iPSCs. Scale bar, 50 μm. (**G**) TEM images of iPSC-derived NPCs. Black arrows indicate mitochondria. Scale bars, 10 μm (*left*) and 500 nm (*right*). (**H**) OCR measurement using iPSC-derived NPCs. (**I**) Quantification of the maximal OCR after the injection of FCCP. Average values ± SEM from three independent experiments were plotted. The value of each replicate was the average calculated from 21 to 40 technical replicate wells. ns, not significant. One-way ANOVA followed by Dunnett's multiple comparisons test.

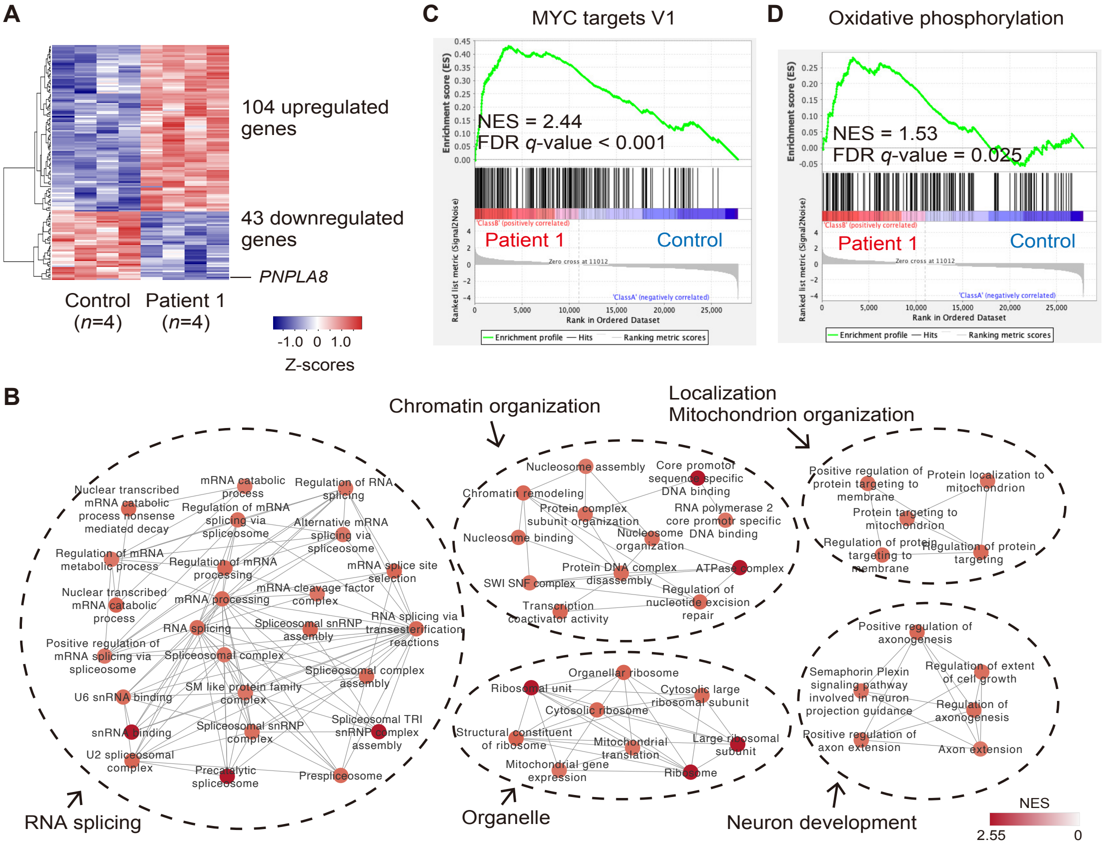


**Supplemental Figure 7. Spatially-resolved differential gene expression related to cell differentiation in cerebral organoids.** (**A**) Heatmaps of the DEGs including 104 upregulated (red) and 43 downregulated (blue) genes in patient aRGCs. (**B**) Enrichment map generated by GSEA tool and Cytoscape Enrichment Map plug-in. Each node represents an enriched GO term. Node color gradient represents normalized enrichment scores (NES). (**C** and **D**) Enrichment plots showing MYC targets V1 (**C**) and oxidative phosphorylation (**D**).

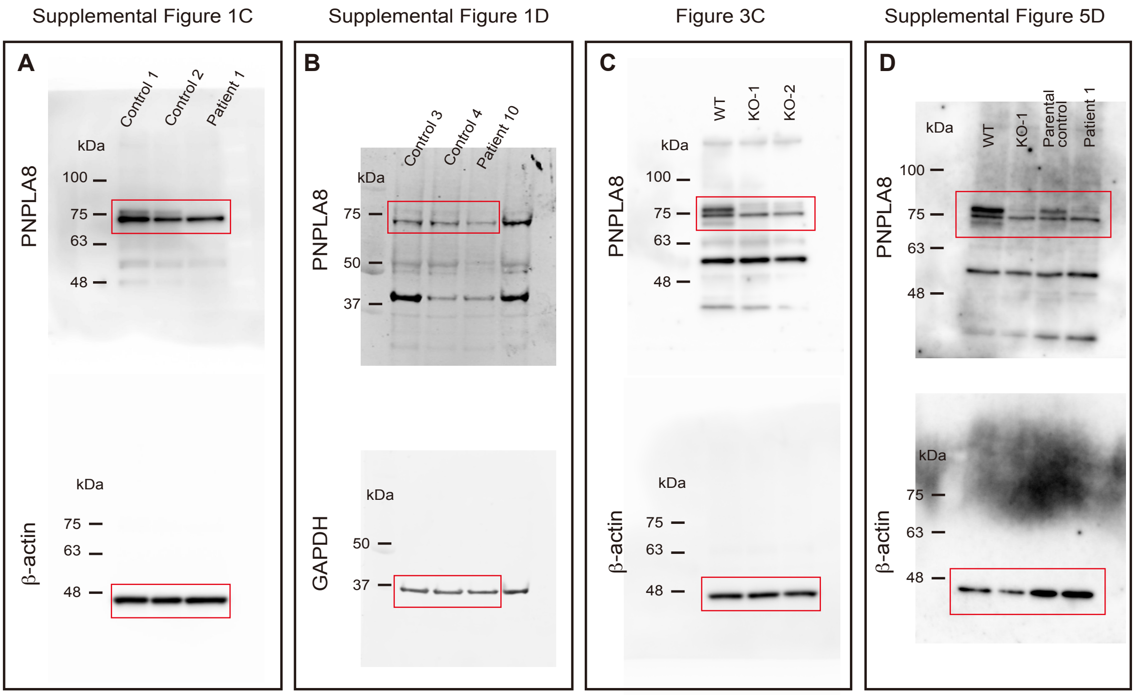


**Supplemental Figure 8. Full-length immunoblot images.** (**A-D**) Full-length immunoblots for Supplemental Figure 1C (**A**), Supplemental Figure 1D (**B**), Figure 3C (**C**), and Supplemental Figure 5D (**D**).

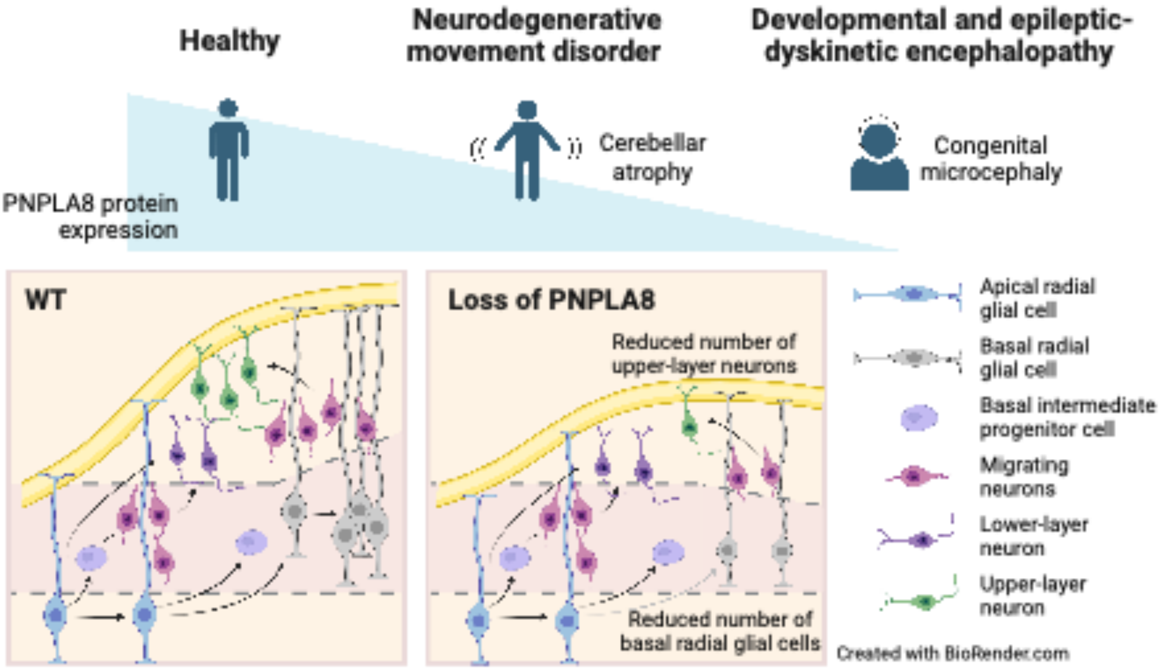


**Graphical abstract**

**Supplemental Table 1.** Additional clinical and genetic details of newly identified individuals with *PNPLA8* variants

| | **Developmental and epileptic-dyskinetic encephalopathy** | | | | | | | **Intermediate** | **Neurodegenerative movement disorder** | | | | **Non-neuronal** | |
|---|---|---|---|---|---|---|---|---|---|---|---|---|---|---|
| Family | 1 | 2 | 3 | 4 | 5 | 6 (Denommé-Pichon *et al.*) | 7 (Harmouch *et al.*) | 8 | 9a | 9b | 9c | 10 | 11 | 12 |
| Gender | Male | Female | Female | Female | Male | Male | Female | Male | Male | Male | Female | Female | Female | Female |
| Age at last assessment (removed due to medRxiv policy) | | | | | | | | | | | | | | |
| Zygosity | CH | CH | Hom | Hom | Hom | Hom | Hom | CH | Hom | Hom | Hom | Hom | Hom | Hom |
| Nucleotide change[a] | c.451A>T/c.1625+3_1625+6del | c.1648del/c.2075-2A>G | c.1748_1749del | c.793dup | c.1684-2A>G | c.944_945del | c.1580G>A | c.1904C>T/c.1637del | c.2275_2276del | c.2275_2276del | c.2275_2276del | c.2275_2276del | c.31_32dup | c.18del |
| Amino acid change | p.Lys151*/[b] | p.Met550*/[b] | p.Tyr583Trpfs*17 | p.Thr265Asnfs*13 | [b] | p.Tyr315* | p.Trp527* | p.Pro635Leu/ p.Gly546Aspfs*4 | p.Leu759Alafs*4 | p.Leu759Alafs*4 | p.Leu759Alafs*4 | p.Leu759Alafs*4 | p.Tyr12Phefs*31 | p.Val7* |
| Gestational period | 34 weeks | Full term | Full term | NA | Full term | Full term | Full term | Full term | NA | NA | NA | Full term | NA | NA |
| Birth HC | 27.8 cm (−2.2 SD) | 28.5 cm (−4.4 SD) | 32.0 cm (−2.0 SD) | NA | NA | 32.0 cm (−1.8 SD) | 28.0 cm (−4.7 SD) | 32 cm (−2.5 SD) | NA | NA | NA | 33.5 (−1.3 SD) | NA | NA |
| Current HC (removed due to medRxiv policy) | 38.6 cm (−6.3 SD) | 32.6 cm (−8.9 SD) | NA | 35.0 cm (−6.6 SD) | 39.0 cm (−6.7 SD) | 39.0 cm (−6.2 SD) | 31.0 cm (−9.4 SD) | 41 cm (−5.1 SD) | NA | NA | NA | 54.0 cm (−0.3 SD) | NA | NA |
| Regression | – | – | – | NA | – | – | – | – | + | + | + | + | – | – |
| Seizure frequency | Intractable, even worse during infection | Intractable | NA | NA | Intractable | Intractable | Intractable | Intractable during infection | Never occurred | One time | Never occurred | Never occurred | Never occurred | Never occurred |
| Ataxia | NA | NA | NA | NA | NA | – | NA | – | + | + | + | + | – | – |
| Myoclonus | – | – | NA | NA | – | – | NA | + | – | + | – | – | – | – |
| Other findings | | | | | | | | | | | | | Deafness | Blindness |

[a]Nucleotide sequences are described with reference to *PNPLA8* transcript NM_001256007.3.
[b]A splicing defect is predicted (see Supplemental Figure 1).
+, present; –, absent; CH, compound heterozygous; d, day; HC, head circumference; Het, heterozygous; Hom, homozygous; mo, month; NA, not available; y, year.

**Supplemental Table 2.** Differentially expressed genes between Control vs Patient 1 cerebral organoids with adjusted *P* values < 0.1.

| Ensemble_gene ID | Gene symbol | Biotype | Chromosome | baseMean | $log_2$ Fold Change (Control vs Patient 1) |
|---|---|---|---|---|---|
| ENSG00000053438 | NNAT | protein_coding | 20 | 423.3410158 | −9.522705541 |
| ENSG00000187653 | TMSB4XP8 | processed_pseudogene | 4 | 16.21993706 | −7.429121365 |
| ENSG00000215319 | EIF5P1 | processed_pseudogene | X | 13.94248912 | −6.467787688 |
| ENSG00000225972 | MTND1P23 | unprocessed_pseudogene | 1 | 1218.314834 | −5.483788946 |
| ENSG00000174080 | CTSF | protein_coding | 11 | 6.75698607 | −5.431715602 |
| ENSG00000158716 | DUSP23 | protein_coding | 1 | 3.644614928 | −5.250650631 |
| ENSG00000274441 | | lncRNA | 5 | 4.399137173 | −4.787397145 |
| ENSG00000274979 | | lncRNA | 12 | 3.36778566 | −4.413834469 |
| ENSG00000248162 | CHORDC1P3 | processed_pseudogene | 4 | 6.845518432 | −3.728887685 |
| ENSG00000243199 | | processed_pseudogene | 4 | 6.430489571 | −3.299220049 |
| ENSG00000187243 | MAGED4B | protein_coding | X | 27.78372823 | −3.127965051 |
| ENSG00000229659 | RPL26P6 | processed_pseudogene | 10 | 17.91758119 | −2.991894355 |
| ENSG00000166770 | ZNF667-AS1 | lncRNA | 19 | 13.60150949 | −2.794050286 |
| ENSG00000247627 | MTND4P12 | processed_pseudogene | 5 | 31.87226585 | −2.238456382 |
| ENSG00000225630 | MTND2P28 | unprocessed_pseudogene | 1 | 88.89615694 | −2.237232779 |
| ENSG00000178464 | RPL10P16 | processed_pseudogene | 19 | 22.65584799 | −2.148183223 |
| ENSG00000162825 | NBPF20 | protein_coding | 1 | 10.18316586 | −2.059293858 |
| ENSG00000180543 | TSPYL5 | protein_coding | 8 | 14.89605299 | −1.966095375 |
| ENSG00000172058 | SERF1A | protein_coding | 5 | 13.81889295 | −1.912859511 |
| ENSG00000105270 | CLIP3 | protein_coding | 19 | 143.6412128 | −1.727852905 |
| ENSG00000234009 | RPL5P34 | processed_pseudogene | 22 | 13.63352778 | −1.715135631 |
| ENSG00000131773 | KHDRBS3 | protein_coding | 8 | 137.2913284 | −1.664754224 |
| ENSG00000225475 | | processed_pseudogene | 1 | 77.34832027 | −1.593292798 |
| ENSG00000184897 | H1-10 | protein_coding | 3 | 58.7457927 | −1.57262416 |
| ENSG00000178057 | NDUFAF3 | protein_coding | 3 | 24.56381569 | −1.50897375 |
| ENSG00000126461 | SCAF1 | protein_coding | 19 | 34.26532737 | −1.485121119 |
| ENSG00000207721 | MIR186 | miRNA | 1 | 17.590077 | −1.438673275 |
| ENSG00000087338 | GMCL1 | protein_coding | 2 | 55.22174563 | −1.37538673 |
| ENSG00000186063 | AIDA | protein_coding | 1 | 28.1240659 | −1.375079975 |
| ENSG00000183666 | GUSBP1 | transcribed_unprocessed_pseudogene | 5 | 27.00796041 | −1.304837766 |
| ENSG00000105669 | COPE | protein_coding | 19 | 33.65480182 | −1.290493491 |
| ENSG00000136295 | TTYH3 | protein_coding | 7 | 80.78316307 | −1.284168833 |
| ENSG00000141698 | NT5C3B | protein_coding | 17 | 31.62712348 | −1.217075781 |
| ENSG00000099942 | CRKL | protein_coding | 22 | 76.56705796 | −1.172980581 |
| ENSG00000118707 | TGIF2 | protein_coding | 20 | 27.1369566 | −1.159105146 |
| ENSG00000100325 | ASCC2 | protein_coding | 22 | 29.47049206 | −1.118027515 |
| ENSG00000142534 | RPS11 | protein_coding | 19 | 692.0368024 | −1.091109054 |
| ENSG00000111676 | ATN1 | protein_coding | 12 | 193.338437 | −1.079751764 |
| ENSG00000263878 | DLGAP1-AS4 | lncRNA | 18 | 63.00604235 | −1.070919253 |
| ENSG00000213315 | | processed_pseudogene | 14 | 44.60211014 | −1.063325048 |
| ENSG00000129559 | NEDD8 | protein_coding | 14 | 34.87036502 | −1.041761822 |
| ENSG00000188612 | SUMO2 | protein_coding | 17 | 385.4735692 | −1.033503067 |
| ENSG00000269028 | MTRNR2L12 | protein_coding | 3 | 76.36786753 | −1.011405507 |
| ENSG00000155980 | KIF5A | protein_coding | 12 | 81.05336185 | −0.98351722 |
| ENSG00000177868 | SVBP | protein_coding | 1 | 89.26164511 | −0.979263308 |
| ENSG00000286259 | | lncRNA | 12 | 69.11582919 | −0.977965576 |
| ENSG00000185591 | SP1 | protein_coding | 12 | 54.5741421 | −0.959345762 |
| ENSG00000126267 | COX6B1 | protein_coding | 19 | 46.44103552 | −0.95820005 |
| ENSG00000179604 | CDC42EP4 | protein_coding | 17 | 76.47542608 | −0.952010265 |
| ENSG00000178531 | CTXN1 | protein_coding | 19 | 179.6629465 | −0.934941061 |
| ENSG00000163041 | H3-3A | protein_coding | 1 | 230.2493182 | −0.910682567 |
| ENSG00000100239 | PPP6R2 | protein_coding | 22 | 57.93524299 | −0.901691178 |
| ENSG00000160200 | CBS | protein_coding | 21 | 83.34275172 | −0.887905581 |
| ENSG00000100823 | APEX1 | protein_coding | 14 | 45.35335014 | −0.879713775 |

| | | | | | |
|---|---|---|---|---|---|
| ENSG00000184939 | ZFP90 | protein_coding | 16 | 63.40165635 | −0.869902674 |
| ENSG00000133627 | ACTR3B | protein_coding | 7 | 51.43944088 | −0.862214826 |
| ENSG00000102226 | USP11 | protein_coding | X | 50.49002754 | −0.853254866 |
| ENSG00000105438 | KDELR1 | protein_coding | 19 | 74.33408717 | −0.836365659 |
| ENSG00000204463 | BAG6 | protein_coding | 6 | 54.57926784 | −0.796202067 |
| ENSG00000213190 | MLLT11 | protein_coding | 1 | 347.1870308 | −0.790105661 |
| ENSG00000087087 | SRRT | protein_coding | 7 | 103.6585649 | −0.761618273 |
| ENSG00000160352 | ZNF714 | protein_coding | 19 | 61.37835543 | −0.758145212 |
| ENSG00000149294 | NCAM1 | protein_coding | 11 | 595.4093304 | −0.750433895 |
| ENSG00000164587 | RPS14 | protein_coding | 5 | 274.4098383 | −0.723736699 |
| ENSG00000099864 | PALM | protein_coding | 19 | 122.7184512 | −0.713684208 |
| ENSG00000100813 | ACIN1 | protein_coding | 14 | 85.8325273 | −0.709907545 |
| ENSG00000174231 | PRPF8 | protein_coding | 17 | 84.3567401 | −0.70529605 |
| ENSG00000110076 | NRXN2 | protein_coding | 11 | 89.85958562 | −0.695823357 |
| ENSG00000166165 | CKB | protein_coding | 14 | 430.8442897 | −0.692410242 |
| ENSG00000212802 | RPL15P3 | processed_pseudogene | 6 | 446.9537437 | −0.675671568 |
| ENSG00000103647 | CORO2B | protein_coding | 15 | 109.4801011 | −0.657677412 |
| ENSG00000180616 | SSTR2 | protein_coding | 17 | 234.7401977 | −0.652734383 |
| ENSG00000249992 | TMEM158 | protein_coding | 3 | 100.731046 | −0.652207829 |
| ENSG00000213923 | CSNK1E | protein_coding | 22 | 207.5750089 | −0.648875984 |
| ENSG00000105576 | TNPO2 | protein_coding | 19 | 144.6368748 | −0.63949019 |
| ENSG00000147677 | EIF3H | protein_coding | 8 | 161.9080355 | −0.638858113 |
| ENSG00000179242 | CDH4 | protein_coding | 20 | 206.053184 | −0.63180802 |
| ENSG00000140988 | RPS2 | protein_coding | 16 | 429.6241168 | −0.61481288 |
| ENSG00000109956 | B3GAT1 | protein_coding | 11 | 197.3755479 | −0.603748078 |
| ENSG00000131116 | ZNF428 | protein_coding | 19 | 146.1433546 | −0.602331245 |
| ENSG00000105968 | H2AZ2 | protein_coding | 7 | 180.4655538 | −0.600991046 |
| ENSG00000076641 | PAG1 | protein_coding | 8 | 140.805984 | −0.595500978 |
| ENSG00000090905 | TNRC6A | protein_coding | 16 | 291.5268044 | −0.574492959 |
| ENSG00000198258 | UBL5 | protein_coding | 19 | 143.3110304 | −0.574404791 |
| ENSG00000109084 | TMEM97 | protein_coding | 17 | 336.161674 | −0.572836823 |
| ENSG00000108671 | PSMD11 | protein_coding | 17 | 179.8372316 | −0.565795213 |
| ENSG00000116350 | SRSF4 | protein_coding | 1 | 138.556805 | −0.555252654 |
| ENSG00000124193 | SRSF6 | protein_coding | 20 | 218.6586562 | −0.555059046 |
| ENSG00000225783 | MIAT | lncRNA | 22 | 339.0637924 | −0.539644194 |
| ENSG00000167658 | EEF2 | protein_coding | 19 | 558.5118102 | −0.529847233 |
| ENSG00000198700 | IPO9 | protein_coding | 1 | 252.5536989 | −0.523788716 |
| ENSG00000186868 | MAPT | protein_coding | 17 | 342.2585511 | −0.523296975 |
| ENSG00000075618 | FSCN1 | protein_coding | 7 | 293.6613276 | −0.519301376 |
| ENSG00000204628 | RACK1 | protein_coding | 5 | 236.0521207 | −0.499785677 |
| ENSG00000167978 | SRRM2 | protein_coding | 16 | 435.9534067 | −0.497482136 |
| ENSG00000135097 | MSI1 | protein_coding | 12 | 255.7857903 | −0.49496585 |
| ENSG00000136631 | VPS45 | protein_coding | 1 | 158.8247764 | −0.493126195 |
| ENSG00000136193 | SCRN1 | protein_coding | 7 | 411.2851068 | −0.490990365 |
| ENSG00000266472 | MRPS21 | protein_coding | 1 | 217.5560133 | −0.490593851 |
| ENSG00000198918 | RPL39 | protein_coding | X | 222.1623549 | −0.486816034 |
| ENSG00000112186 | CAP2 | protein_coding | 6 | 239.8469967 | −0.485847242 |
| ENSG00000170004 | CHD3 | protein_coding | 17 | 295.4346219 | −0.481026243 |
| ENSG00000205542 | TMSB4X | protein_coding | X | 1138.558977 | −0.425057748 |
| ENSG00000148798 | INA | protein_coding | 10 | 591.9931424 | −0.38131134 |
| ENSG00000166257 | SCN3B | protein_coding | 11 | 553.0871187 | 0.38813213 |
| ENSG00000196591 | HDAC2 | protein_coding | 6 | 697.4370379 | 0.407549592 |
| ENSG00000139970 | RTN1 | protein_coding | 14 | 639.7187188 | 0.411999178 |
| ENSG00000119185 | ITGB1BP1 | protein_coding | 2 | 228.9759316 | 0.484159589 |
| ENSG00000131174 | COX7B | protein_coding | X | 445.6025658 | 0.494218615 |
| ENSG00000185088 | RPS27L | protein_coding | 15 | 381.6898085 | 0.557750138 |
| ENSG00000169180 | XPO6 | protein_coding | 16 | 135.8819104 | 0.597586095 |
| ENSG00000089818 | NECAP1 | protein_coding | 12 | 142.7802307 | 0.672978315 |

| | | | | | |
|---|---|---|---|---|---|
| ENSG00000129355 | CDKN2D | protein_coding | 19 | 179.9878323 | 0.673415346 |
| ENSG00000169760 | NLGN1 | protein_coding | 3 | 81.92803571 | 0.694608933 |
| ENSG00000196428 | TSC22D2 | protein_coding | 3 | 99.82182844 | 0.700601118 |
| ENSG00000169744 | LDB2 | protein_coding | 4 | 123.8186651 | 0.704904803 |
| ENSG00000083720 | OXCT1 | protein_coding | 5 | 82.31067179 | 0.706616806 |
| ENSG00000115461 | IGFBP5 | protein_coding | 2 | 697.5810285 | 0.756291397 |
| ENSG00000135241 | PNPLA8 | protein_coding | 7 | 100.9338426 | 0.855986806 |
| ENSG00000281344 | HELLPAR | lncRNA | 12 | 67.14649707 | 0.876567768 |
| ENSG00000197301 | HMGA2-AS1 | lncRNA | 12 | 89.03080685 | 0.884117081 |
| ENSG00000213420 | GPC2 | protein_coding | 7 | 92.76489775 | 0.891322807 |
| ENSG00000157168 | NRG1 | protein_coding | 8 | 88.80996985 | 0.902306484 |
| ENSG00000158470 | B4GALT5 | protein_coding | 20 | 299.6149984 | 0.933661035 |
| ENSG00000130066 | SAT1 | protein_coding | X | 56.86521824 | 1.068842339 |
| ENSG00000234498 | RPL13AP20 | processed_pseudogene | 12 | 30.45043018 | 1.13805603 |
| ENSG00000156140 | ADAMTS3 | protein_coding | 4 | 35.23930563 | 1.195621364 |
| ENSG00000197728 | RPS26 | protein_coding | 12 | 44.37274857 | 1.290626368 |
| ENSG00000213965 | NUDT19 | protein_coding | 19 | 21.21645532 | 1.294153907 |
| ENSG00000233762 | RPS15P4 | processed_pseudogene | 2 | 131.4383563 | 1.41060517 |
| ENSG00000241743 | XACT | lncRNA | X | 58.84892991 | 1.411662913 |
| ENSG00000187824 | TMEM220 | protein_coding | 17 | 20.28981761 | 1.479002366 |
| ENSG00000248527 | MTATP6P1 | unprocessed_pseudogene | 1 | 158.7276468 | 1.534083015 |
| ENSG00000164176 | EDIL3 | protein_coding | 5 | 81.63041546 | 1.55732981 |
| ENSG00000278532 | | lncRNA | 18 | 34.34571532 | 1.557492908 |
| ENSG00000277586 | NEFL | protein_coding | 8 | 18.57903878 | 1.629604163 |
| ENSG00000279090 | | TEC | 11 | 25.34535404 | 1.698866137 |
| ENSG00000124762 | CDKN1A | protein_coding | 6 | 55.80704502 | 1.797151729 |
| ENSG00000244694 | PTCHD4 | protein_coding | 6 | 20.12130454 | 2.131201605 |
| ENSG00000226752 | CUTALP | transcribed_unitary_pseudogene | 9 | 10.31467338 | 2.513686868 |
| ENSG00000170927 | PKHD1 | protein_coding | 6 | 6.273646181 | 2.573625984 |
| ENSG00000180229 | HERC2P3 | transcribed_unprocessed_pseudogene | 15 | 55.84801905 | 2.92323542 |
| ENSG00000131409 | LRRC4B | protein_coding | 19 | 7.396233402 | 3.525418038 |
| ENSG00000230383 | | processed_pseudogene | 7 | 7.365153954 | 4.278613716 |
| ENSG00000267890 | | lncRNA | 19 | 8.71829341 | 4.588856367 |
| ENSG00000283930 | PLD5P1 | protein_coding | 10 | 14.6476857 | 4.636562379 |
| ENSG00000184110 | EIF3C | protein_coding | 16 | 27.67850074 | 4.68427797 |

**Supplemental Table 3.** Oligonucleotides, antibodies, plasmids, and reagents used in this study.

| **Oligonucleotides** | **Source** | **Sequences (5' to 3')** |
|---|---|---|
| RT-PCR and Sanger sequencing for exons 3 to 9 of *PNPLA8* (Forward) | Eurofins Genomics | AGGCTGTTTTTGGCAATC |
| RT-PCR and Sanger sequencing for exons 3 to 9 of *PNPLA8* (Reverse) | Eurofins Genomics | CACTCTAACGGCACATCT |
| PNPLA8 sgRNA-1 (Design ID: Hs.Cas9.PNPLA8.1.AA) | Integrated DNA Technologies | GGTATCGCAGAAGGACTTGT |
| PNPLA8 sgRNA-2 (Design ID: Hs.Cas9.PNPLA8.1.AK) | Integrated DNA Technologies | CAAGGGTGAGTATTGATAAC |
| **Antibodies** | **Source** | **Identifier** |
| SOX17 | R and D Systems | Cat.# AF1924 |
| αSMA | Sigma-Aldrich | Cat.# A2547 |
| TUJ1 (Supplemental Figure 5B) | Sigma-Aldrich | Cat.# T8660 |
| PNPLA8 | Atlas Antibodies | Cat.# HPA020083 |
| β-actin | MBL | Cat.# M177-3 |
| GAPDH | Thermo Fisher Scientific | Cat.# AM4300 |
| SOX2 | Santa Cruz | Cat.# sc-365823 |
| TBR2 | Abcam | Cat.# ab275960 |
| CTIP2 | Abcam | Cat.# ab18465 |
| SATB2 | Santa Cruz | Cat.# sc-81376 |
| TUJ1 (Supplemental Figure 3A) | Santa Cruz | Cat.# sc-80005 |
| PAX6 | BioLegend | Cat.# 901301 |
| HOPX | Santa Cruz | Cat.# sc-398703 |
| BrdU | Abcam | Cat.# ab6326 |
| Ki67 | Abcam | Cat.# ab16667 |
| pH3 | Santa Cruz | Cat.# sc-374669 |
| Cleaved Caspase 3 | CST | Cat.# 9661S |
| NESTIN | Santa Cruz | Cat.# sc-23927 |
| Anti-Mouse IgG-Peroxidase antibody | Sigma-Aldrich | Cat.# A9044 |
| Anti-Rabbit IgG–Peroxidase antibody | Sigma-Aldrich | Cat.# A0545 |
| IRDye 800CW Goat anti-Mouse IgG | Li-cor Biosciences | Cat.# 926-32210 |
| IRDye 800CW Goat anti-Rabbit IgG | Li-cor Biosciences | Cat.# 926-32211 |
| Goat anti-Rabbit Secondary Antibody, Alexa Fluor™ 488 | Invitrogen | Cat.# A-11008 |
| Goat anti-Mouse IgG2b Secondary Antibody, Alexa Fluor™ 488 | Invitrogen | Cat.# A-21141 |
| Goat anti-Rabbit Secondary Antibody, Alexa Fluor™ 594 | Invitrogen | Cat.# A-11037 |
| Goat anti-Rat Secondary Antibody, Alexa Fluor™ 555 | Invitrogen | Cat.# A-21434 |
| Goat anti-Mouse IgG2a Secondary Antibody, Alexa Fluor™ 555 | Invitrogen | Cat.# A-21137 |
| Goat anti-Mouse IgG1 Secondary Antibody, Alexa Fluor™ 647 | Invitrogen | Cat.# A-21240 |
| **Plasmids** | **Source** | **Identifier** |
| pCE-hOCT3/4 | Addgene | Cat.# 41813 |
| pCE-hSK | Addgene | Cat.# 41814 |
| pCE-hUL | Addgene | Cat.# 41855 |
| pCE-mp53DD | Addgene | Cat.# 41856 |
| pCXB-EBNA1 | Addgene | Cat.# 41857 |
| **Reagents** | **Source** | **Identifier** |
| QIAamp DNA Blood Midi Kit | Qiagen | Cat.# 51185 |
| Sureselect XT Human All Exon V6 capture library | Agilent Technologies | Cat.# 5190-8863 |
| Cyclosporin A | Sigma-Aldrich | Cat.# SML1018 |
| RPMI-1640 | Sigma-Aldrich | Cat.# R6504 |
| Fetal Bovine Serum (FBS) | Gibco | Cat.# 16000044 |
| L-Glutamine–Penicillin–Streptomycin solution | Sigma-Aldrich | Cat.# G1146 |

| | | |
|---|---|---|
| Dulbecco's Modified Eagle Medium (DMEM) | Gibco | Cat.# C11995500 |
| Normal human dermal fibroblasts | PromoCell | Cat.# C-12300 |
| RNeasy mini kit | Qiagen | Cat.# 74106 |
| DNA-free™ DNA Removal Kit | Thermo Fisher Scientific | Cat.# AM1906 |
| High-Capacity cDNA Reverse Transcription Kit | Thermo Fisher Scientific | Cat.# 4368814 |
| TaqMan™ Fast Advanced Master Mix | Thermo Fisher Scientific | Cat.# 4444556 |
| SuperScript IV Reverse Transcriptase | Invitrogen | Cat.# 18090010 |
| AmpliTaq Gold 360 Master Mix | Applied Biosystems | Cat.# 4398881 |
| TOPO TA Cloning kit | Invitrogen | Cat.# 450641 |
| One Shot TOP10 Chemically Competent E. coli | Invitrogen | Cat.# C404010 |
| QIAprep Spin Miniprep Kit | Qiagen | Cat.# 27106 |
| PBS (-) | Wako | Cat.# 166-23555 |
| Sodium Dodecyl Sulfate | Wako | Cat.# 199-07141 |
| Tris–HCl | NIPPON GENE | Cat.# 318-90225 |
| Glycerol | Wako | Cat.# 075-00616 |
| 2-mercaptoethanol | Gibco | Cat.# 21985-023 |
| Bromophenol blue | Wako | Cat.# 029-02912 |
| RIPA lysis buffer | Wako | Cat.# 182-02451 |
| Protease inhibitor cocktail | Roche | Cat.# 11836153001 |
| e-PAGEL (7.5%) | ATTO Corporation | Cat.# 2331800 |
| Polyvinylidene difluoride membrane | Millipore | Cat.# IPVH00010 |
| WIDE-VIEW™ Western Protein Size Marker III | Wako | Cat.# 234-02464 |
| ECL™ Prime Western Blotting Detection Reagent | Cytiva | Cat.# RPN2236 |
| Western BLoT Stripping Buffer | Takara Bio | Cat.# T7135A |
| Nitrocellulose Membrane | Bio-Rad | Cat.# 1620115 |
| Glutaraldehyde Solution | Electron Microscopy Sciences | Cat.# 111-30-8 |
| Osmium tetroxide | Sigma-Aldrich | Cat.# 1.24505 |
| Quetol 812 epoxy resin | Nissin | Cat.# 340-H |
| Uranyl acetate | Merck | Cat.# 8473 |
| Seahorse XFe96/XF Pro FluxPak Mini | Agilent | Cat.# 103793-100 |
| Seahorse XF DMEM Medium | Agilent | Cat.# 103575-100 |
| CyQUANT Cell Proliferation Assay Kit | Invitrogen | Cat.# C7026 |
| Seahorse XF Cell Mito Stress Test Kit | Agilent | Cat.# 103015-100 |
| Seahorse XF 1.0M Glucose solution | Agilent | Cat.# 103577-100 |
| Seahorse XF 100mM Pyruvate solution | Agilent | Cat.# 103578-100 |
| Seahorse XF 200mM Glutamine solution | Agilent | Cat.# 103579-100 |
| StemFit® AK02N | Ajinomoto | Cat.# RCAK02N |
| Y-27632 | Sigma-Aldrich | Cat.# 688000 |
| iMatrix-511 (recombinant laminin-511) | Takara Bio | Cat.# T304 |
| Accutase | Nacalai Tesque | Cat.# 12679-54 |
| tracrRNA with Atto550 | Integrated DNA technologies | Cat.# 1075927 |
| HiFi Cas9 Nuclease V3 | Integrated DNA technologies | Cat.# 1081060 |
| Opti-MEM™ I Reduced Serum Medium | Gibco | Cat.# 31985062 |
| StemSpan™ H3000 | STEMCELL Technologies | Cat.# 09850 |
| Recombinant human interleukin-6 | Pepro Tech | Cat.# 200-06 |
| Recombinant human stem cell factor | Pepro Tech | Cat.# 300-07 |
| Recombinant human thrombopoietin | Pepro Tech | Cat.# 300-18 |
| Recombinant human Flt3 ligand | Pepro Tech | Cat.# 300-19 |
| Recombinant human interleukin-3 | Pepro Tech | Cat.# 200-03 |
| Human CD34+ Cell Nucleofector Kit | Lonza | Cat.# VPA-1003 |
| Mitomycin C | Sigma-Aldrich | Cat.# M4287 |
| Nunclon Sphera 96V plate | Thermo Fisher Scientific | Cat.# 81100675 |

| | | |
|---|---|---|
| DMEM/F12 | Invitrogen | Cat.# 11330-032 |
| KnockOut Serum Replacement | Thermo Fisher Scientific | Cat.# 10828010 |
| MEM Non-essential amino acids solution (100X) | Gibco | Cat.# 11140 |
| L-glutamine | Thermo Fisher Scientific | Cat.# 25030081 |
| LDN-193189 | Cayman Chemical | Cat.# 19396 |
| SB-431542 | Cayman Chemical | Cat.# 13031 |
| XAV939 | Cayman Chemical | Cat.# 13596 |
| 2-mercaptoethanol | Wako | Cat.# 133-14571 |
| 2-hydroxyethyl methacrylate | Sigma-Aldrich | Cat.# P3932 |
| MACS Neuro Medium | Miltenyi Biotec | Cat.# 130-093-570 |
| Insulin | Nacalai Tesque | Cat.# 12878-86 |
| N2 supplement | R and D Systems | Cat.# AR009 |
| CnT Pen-Strep / Amphotericin B Solution, 250 x, Ready-to-Use Single Aliquots | CellnTES | Cat.# Cnt-ABM10 |
| NeuroBrew-21 supplement without VitA | Miltenyi Biotec | Cat.# 130-097-263 |
| NeuroBrew-21 supplement | Miltenyi Biotec | Cat.# 130-093-566 |
| L-Ascorbic acid | Sigma-Aldrich | Cat.# A92902 |
| Disposable surgical scalpel | Kai medical | Cat.# No. 10 510-A |
| EZ-sphere 24-well plates | Iwaki | Cat.# 4820-900SP |
| Matrigel | Corning | Cat.# 354234 |
| Triton X-100 | Sigma-Aldrich | Cat.# X100 |
| Blocking-One | Nacalai Tesque | Cat.# 03953-95 |
| Fluoromount | Diagnostic BioSystems | Cat.# K 024 |
| Poly-L-Lysine | Sigma-Aldrich | Cat.# P8920 |
| Laminin Mouse Protein, Natural | Invitrogen | Cat.# 23017-015 |
| Recombinant human EGF protein | Almone Labs | Cat.# E-100 |
| Stem Cell Banker | Takara | Cat.# CB045 |
| 96-well glass bottom plate | Matsunami | Cat.# GP96000 |
| Paraformaldehyde | Wako | Cat.# 162-16065 |
| Sucrose | Nacalai Tesque | Cat.# 30403-55 |
| Tissue-Tek O.C.T. Compound | Sakura Finetek Japan | Cat.# 4583 |
| DAPI | Cayman Chemical | Cat.# 14285 |
| Fluoromount-G | SouthernBiotech | Cat.# 0100-01 |
| 5-Bromo-2′-deoxyuridine | Sigma-Aldrich | Cat.# B5002 |
| Hydrochloric acid | Sigma-Aldrich | Cat.# 13-1640 |
| Proteinase K | Kanto Chemical | Cat.# 34060-96 |
| MEGAscript™ T7 Transcription Kit | Invitrogen | Cat.# AMB13345 |
| 2-Methylbutane | Wako | Cat.# 166-00615 |
| Cardiolipin 56:0 | Avanti Polar Lipids | Cat.# 750332 |
| Phosphatidylcholine 25:0 | Avanti Polar Lipids | Cat.# LM1000 |
| Phosphatidylethanolamine 25:0 | Avanti Polar Lipids | Cat.# LM1100 |
| Phosphatidylglycerol 25:0 | Avanti Polar Lipids | Cat.# LM1200 |
| Phosphatidylinositol 25:0 | Avanti Polar Lipids | Cat.# LM1500 |
| Phosphatidylserine 25:0 | Avanti Polar Lipids | Cat.# LM1300 |
| Phosphatidic acid 34:0 | Avanti Polar Lipids | Cat.# 830856 |
| Lysophosphatidylcholine-d49 16:0 | Avanti Polar Lipids | Cat.# 870308 |
| Lysophosphatidyletanolamine-d7 18:1 | Avanti Polar Lipids | Cat.# 791644 |
| Lysophosphatidylglycerol 17:1 | Avanti Polar Lipids | Cat.# 858127 |
| Lysophosphatidylinositol 17:1 | Avanti Polar Lipids | Cat.# 850103 |
| Lysophosphatidylserine 17:1 | Avanti Polar Lipids | Cat.# 858141 |
| Lysophosphatidic acid 17:0 | Avanti Polar Lipids | Cat.# 857127 |
| Arachidonic acid-d8 | Cayman Chemical | Cat.# AA-d8 |
| DC™ Protein Assay Kit I | Bio-Rad | Cat.# 5000111 |

| | | |
|---|---|---|
| Chloroform | Merck | Cat.# 103420 |
| Ammonium formate | Merck | Cat.# 70221 |
| Formic acid | Merck | Cat.# 100264 |
| (C14:0)4 cardiolipin sodium salt | Avanti Polar Lipids | Cat.# 750332 |
